# Assessing Foundation Models for Computational Pathology in Endometrial Cancer

**DOI:** 10.64898/2026.05.22.26353897

**Authors:** Sarah Volinsky-Fremond, Nikki van den Berg, Jurriaan Barkey Wolf, Lydia A. Schoenpflug, Sonali Andani, Gitte Ørtoft, Jan J. Jobsen, Ludy C. H. W. Lutgens, Melanie E. Powell, Linda R. Mileshkin, Helen Mackay, Alexandra Leary, Rubina R. Razack, Marco de Bruyn, Stephanie M. de Boer, Remi A. Nout, Vincent T. H. B. M. Smit, Carien L. Creutzberg, Viktor H. Koelzer, Tjalling Bosse, Nanda Horeweg

## Abstract

Computational pathology leverages deep learning to extract clinically relevant information from digitized tumor slides, predicting histopathological subtypes, molecular alterations, and patient outcomes. Recent pipelines increasingly rely on foundation models trained on large pan-cancer datasets to generate generalizable features. In endometrial cancer (EC), their comparative performance for clinical diagnostic tasks remains unexplored. For the first time, this study evaluates the performance of seven state-of-the-art foundation models across morphological, molecular, and prognostic tasks using a large EC dataset of 3,293 patients from randomized trials and clinical cohorts. In addition, their performance was compared to one model (EsVIT) exclusively trained on EC. The foundation models H-OPTIMUS-0, CONCH, and VIRCHOW2, achieved the highest mean performance, but the best-performing foundation model varied by task. The top-performing foundation model outperformed the EC-specific feature extractor EsVIT across all tasks. This study highlights the superiority of foundation models over a domain-specific feature extractor in EC. Selecting the optimal foundation model for novel tasks remains challenging due to performance plateaus and limited information on the training datasets, requiring rigorous benchmarking and domain insight to reach maximum potential.

## Introduction

Histopathological evaluation of tumor grade, histologic subtype, and molecular alterations is essential for the diagnosis and accurate risk stratification of malignant tumors. Complete and accurate classification often requires ancillary tests, such as immunohistochemistry or molecular sequencing, which increase diagnostic time, complexity, and costs. With rising cancer incidence and growing workloads for pathologists, there is a need for integrative, high-performing support tools. Artificial intelligence (AI) has the potential to complement pathologists by providing automated assessment of histologic features, tumor grade, and biomarker status, making workflows more efficient and reducing variability.

Computational pathology, particularly deep learning (DL), enables automated extraction of clinically meaningful information from digitized whole-slide images (WSIs). Across multiple cancer types, DL models have demonstrated the ability to predict histologic subtyping and tumor grades^1–8^, molecular alterations^9–17^ and patient outcomes^18–23^. In endometrial cancer (EC), studies have shown that DL can classify the four molecular subtypes^9^, perform histologic grading^24^, and estimate recurrence risk^18^. These advances illustrate the potential of AI to augment routine pathology workflows and become an integral component of diagnostic pathology.

A key component in the DL pipelines for such computational pathology tasks is feature extraction from WSIs. These high resolution images are too large in size for direct processing, and are therefore decomposed into smaller regions, from which compact and informative feature embeddings are derived. Traditionally, feature extraction relied on models pretrained on arbitrary images that vary in shape and color (e.g., ImageNet^25^), but these models are suboptimal for histopathology. Recently, very large DL models trained on pan-cancer images have been proposed, also known as foundation models.^26–31^ Foundation models encode high-resolution tiles into information-rich representations that generalize across tasks and cancer types. As such, foundation models can enhance predictive performance while reducing the need for task-specific retraining.

Benchmarking studies across multiple cancers have consistently shown a high performance of foundation models, such as VIRCHOW2, UNI(-v2), H-OPTIMUS-0(/-1), and CONCH.^27,29,32,33^ Nonetheless, selecting an appropriate foundation model for a specific disease or task is challenging because there are few studies investigating performance for cancer-specific applications. An example is Breen et al.’s assessment of 14 foundation models for ovarian cancer subtyping, demonstrating that cancer-specific benchmarking can provide additional relevant insights.^34^ In endometrial cancer, no such study has been performed to our knowledge.

An alternative for using foundation models is to use feature encoders trained exclusively on cancer-specific images. For example, when Volinsky-Fremond et al. predicted distance recurrence in EC, an EC-specific encoder outperformed at the time state-of-the-art foundation model CTransPath.^18,35^ Interestingly, the EC-specific encoder was trained on three million EC tiles from 1,800 patients, while CTransPath was trained on 15 million tiles from 12,000 patients. This may indicate that pan-cancer models underrepresent subtle, cancer-specific morphological patterns. No comparative study on the performance of foundation models next to EC-specific encoder models in EC has been conducted to date, likely due to the scarcity of large, well-annotated datasets.

To address this gap, we present the first systematic benchmark of state-of-the-art foundation models in EC. For this, we use a large EC dataset, comprising 3,293 patients from three phase III randomized trials, one phase II trial, and eight additional clinical cohorts. We evaluate seven foundation models across eight clinically relevant tasks, including morphological subtyping, detection of molecular features, and prediction of recurrence. We also compare these models to a domain-specific EC feature extractor trained solely on EC data. This study is the first to provide guidance for selecting foundation models in endometrial cancer, supporting the development of AI-based diagnostic tools that enhance pathology workflows.

## Materials and methods

### Ethics

The PORTEC-1 (no ethics registration number, took place in 90s), PORTEC-2 (P01.146), and PORTEC-3 (P06.031), DOMEC study protocols were approved by the Medical Ethical Committee Leiden, Den Haag, Delft and the medical ethics committees at participating centers. All study participants of the clinical trials provided informed consent. Studies were conducted in accordance with the principles of the Declaration of Helsinki. Permissions for the retrospective use of the cohorts and a waiver for informed consent was provided for the Transportec High Risk Pilot (B21.065), Medisch Spectrum Twente cohorts (MST, B21.011), Leiden cohort (nWMO-D4-2023-002), and South Africa cohort (HREC: N23/09/119). Permission for use of the Danish Cohort was provided by the Center for Regional Udvikling, De Videnskabsetiske Komiteer (H-16025909). For the UMCG cohort, the medical ethical committee granted permission for the use of the data and provided a waiver for informed consent owing to the observational nature of the study.

### Trial registration

PORTEC 1: Clinical trial number: not applicable. PORTEC-2 is registered in ISRCTN, number: ISRCTN16228756, date: 20/12/2005. PORTEC-3 is registered in ISRCTN, number: ISRCTN14387080, date: 28/09/2006. DOMEC is registered at ClinicalTrials.gov, number: NCT03951415, date: 01/04/2021.

### Cohorts

We used formalin-fixed paraffin-embedded (FFPE) tumor material and clinicopathological data of patients with EC from 12 cohorts: the PORTEC-1 randomized trial^36^ the PORTEC-2 randomized trial^37^, the PORTEC-3 randomized trial^38^, the DOMEC phase II trial^39^, the Danish cohort^40^, two prospective cohorts of the Medisch Spectrum Twente (MST-I^41^ and MST-II)^42^, the transPORTEC study^43^, the Leiden University Medical Center (LUMC) cohort^18^, the University Medical Center Groningen (UMCG) cohort^44^ the South-Africa cohort^45^, and the publicly available TCGA-UCEC^46^.

The PORTEC-1 randomized trial, conducted between 1990 and 1997, enrolled 714 women with early-stage, intermediate-risk EC to receive either pelvic external beam radiotherapy or no adjuvant therapy. The PORTEC-2 trial, carried out from 2000 to 2006, included 427 women with early-stage, high-intermediate-risk EC, who were randomized to external beam radiotherapy or vaginal brachytherapy. The PORTEC-3 trial recruited 660 women with stage I-III high-risk EC between 2006 and 2013, to receive pelvic external beam radiotherapy alone or external beam radiotherapy combined with concurrent and adjuvant chemotherapy. The DOMEC is a multicenter phase II trial investigating PD-L1 and PARP inhibition for advanced EC and enrolled 55 metastatic or recurrent patients with EC from July 2019 to November 2020. The Danish cohort included 451 grade 3 EC cases registered prospectively in the Danish gynecological cancer database. The prospective MST-I cohort included 257 patients with stage I-III high-risk EC using the same criteria as PORTEC-3, treated between 1987 and 2015 at MST in the Netherlands. Similarly, the MST-II cohort comprised 173 patients with stage I intermediate/high-intermediate-risk EC treated during the same timeframe at MST Enschede. The retrospective TransPORTEC study analyzed 116 high-risk EC tumors collected from five international institutions: Leiden University Medical Center (The Netherlands), University Medical Center Groningen (The Netherlands), University College London (United Kingdom), St. Mary’s Hospital in Manchester (United Kingdom), and Gustave Roussy Institute in Villejuif (France). The LUMC cohort consisted of 222 patients diagnosed and treated at Leiden University Medical Center from 2012 to 2021. The UMCG cohort included 278 patients treated at the University Medical Center Groningen between 1984 and 2004, with follow-up data extending to 2010. The South-Africa cohort is a retrospectively collected high-risk population of 150 patients with EC, from January 2017 to December 2021, diagnosed and treated at Tygerberg Academic Hospital (Cape Town, South Africa). Lastly, the TCGA-UCEC cohort of 529 patients was accessed via the cBioPortal public database^46^.

### Dataset

Only patients with one digitized H&E-stained tumor slide were included. Next, to avoid any overlap between the patch-level feature extractor and the supervised tasks, the 1,842 patients of pooled cohort^18^ (which included the TCGA-UCEC) who were already used for training the patch-level feature extractor model with SSL were excluded for supervised training. Across the foundation models, only the publicly available TCGA-UCEC was used for training. As a result, the final dataset of pooled cohorts that can be used for supervised training included 1,451 patients (with one H&E-stained WSI): 120 from the PORTEC-1 randomized trial, 100 from the PORTEC-2 randomized trial, 268 from the PORTEC-3 randomized trial, 45 from the DOMEC trial, 142 from the DANISH cohort, 52 from MST-I, 174 MST-II, 28 from the transPORTEC study, 203 from LUMC cohort, 177 from the UMCG cohort, 142 from the SOUTH-AFRICA cohort.

All patients were randomly allocated to one of five folds to implement a five-fold cross-validation routine stratified by the ground-truth label. In the case of the distant recurrence task, the stratification was performed on the censorship status and the time bins.

### Supervised tasks

To compare the foundation models and SSL models across relevant EC-specific tasks, the supervised tasks covered three morphology tasks, four molecular tasks, and one prognostic task. All models for the tasks were trained and predicted using H&E-stained WSIs digitized at 40x magnification. For each, patients will be excluded when the ground-truth label is not available or unknown (Supplementary Tables S1-S8).

The first morphology task was classifying patients as low-grade (pooled grade 1 and grade 2) endometrioid (EEC) or high-grade EEC using 1,036 patients (736 low-grade EEC and 300 high-grade EEC). The second morphology task was EEC versus non-EEC (pooled serous, clear cell, mixed, ambiguous, carcinosarcoma, de/di-differentiated) with a total number of 1,436 patients (1,036 EEC and 400 non-EEC). Subsequently, the same task was complexified into a three-class histological subtyping: EEC versus serous (SEC) versus clear cell (CCC), with a total number of 1,310 cases, of which 1,036 were EEC, 202 were SEC, and 72 were CCC.

The four molecular tasks were binary and included 1) *POLE* mutant (*POLE*mut) versus *POLE* wildtype with 1,026 patients (of which 81 were *POLE*mut EC); 2) MMRd versus MMRp with 1,137 patients (of which 306 were MMRd); and 3) p53 abnormal versus wildtype with 1,125 patients (of which 265 were abnormal); 4) ER positive versus ER negative with 1,152 patients (of which 884 is ER positive). *POLE* mutational ground truth data was obtained with DNA sequencing (NGS). Mutant-type staining was used to assign cases as p53abn EC, or by NGS (including the TP53 gene) when IHC failed or was ambiguous. MMRd status was assessed with IHC for the four or the two MMR antibody approach^47^. ER positivity was assessed with IHC, and a cutoff of 10% was used for positivity^41^. For the purpose of this study, we use the IHC ground truth rather than the WHO 2020 diagnostic algorithm of EC^48^, which disregards MMR deficiency and p53 abnormality when the patient is *POLE*mut, and which disregards p53 abnormality when the patient is MMRd (and *POLE* wildtype).

The prognostic task was defined as predicting the distant-recurrence-free probability and included 849 patients (six-year median follow-up; 112 events). Patients with stage IV or those who underwent adjuvant chemotherapy were excluded. Time was binarized into 4 bins following the distribution of uncensored patients for training the attention multiple instance learning model with a batch size of 1^18^.

### Feature extractor models

Seven state-of-the-art patch-level foundation models for which weights are publicly available were used: H-OPTIMUS-0^32^; GIGAPATH^30^; VIRCHOW2^29^; HIBOU-L^31^; UNI^27^; CONCH^26^; CTRANSPATH^35^.

H-OPTIMUS-0 produces 1536-long patch features using a 1.1B parameter ViT-G (‘giant’) architecture, trained on 273M tiles from 500,000 H&E-stained WSIs at 20× magnification using the DINOv2^32^ training scheme. The number of patients with EC included in the training dataset was not disclosed.

GIGAPATH produces 1536-long patch features using a 1.1B parameter ViT-G architecture, trained on 1.3M tiles from 171,189 WSIs of 30,000 patients at 20× magnification with the DINOv2 training scheme. WSIs were both H&E and IHC-stained. The training dataset included 1,847 EC WSIs.

VIRCHOW2 produces a 2560-long patch feature vector (stacking the 1280-long class token and the averaged 1280-long patch tokens) using a 632M parameter ViT-H (huge) architecture trained on 1.9B tiles from 1.5M H&E-stained WSIs of 120,000 patients using the DINOv2 training scheme. The training dataset included 50,610 EC WSIs.

HIBOU-L (Large) produces a 1024-long patch feature vector using a 307M parameter ViT-L (large) architecture, trained on 1.2B tiles from 1.1M WSIs of 306,000 patients using the DINOv2 training scheme. H&E-stained, IHC-stained, and cytology images were included. The number of patients with EC in the training dataset was not disclosed.

UNI produces a 1024-long patch feature vector using a 307M parameter ViT-L architecture, trained on 100M tiles from 100,400 H&E-stained WSIs (the number of patients was not disclosed) using the DINOv2 training scheme. H&E-stained, non-H&E, and cytology images were included. 243 slides with EC were included.

CONCH is a multimodal model producing 512-long patch features from a 90M-parameter ViT-B (big) image encoder. It was trained using 1.17M histology image–caption pairs and the CoCa routine scheme. The number of images showing EC in the training dataset was not disclosed in the publication.

CTRANSPATH produces a 768-long patch feature using a 28M parameter Swin-T (tiny) architecture with a window size of 14 and the MoCo-v3 training scheme. It was trained on 15M tiles from 32,000 H&E-stained WSIs of 12,000 patients. The TCGA and PAIP datasets were used for training; hence, we estimate around 500 slides of EC.

EsVIT^49^ with a modified architecture of 68M parameters was trained in a previous publication^18^. The architecture employed a window size of 14, a patch of 8, and a Swin-T architecture. It was trained with the MoCo-v3 training scheme. The training dataset comprised 3.7M of EC tiles from 1,837 EC patients exhibiting different morphology (55% low-grade EEC, 26% high-grade EEC, 11% serous, and 8% other non-EEC subtypes). Extracting the last block produces 768-long features, and the last 8 blocks 3,456-long features.

### Supervised training

All task-specific datasets were split following a five-fold cross-validation scheme stratified by the supervised label. In the case of the prognostic task, the split was based on both the censorship status and the time bin. Given the patch-level features, an attention multiple instance learning model was trained and optimized on each fold, and performance on each fold was reported with the best set of hyperparameters on average. All classification tasks were optimized based on the AUC, and the prognostic task one based on the C-index metric^50^ (using a tau = 10 years and scikit-survival Python package (version 0.17.2)).

The attention multiple instance learning model comprised an attention module that compressed the feature size to 256 and encoded the patient-level embedding into the feature size of the patch (output of the patch encoder). One extra fully connected layer was added to compress the slide-level embedding to 128, followed by the task head. The AdamW optimizer with its Pytorch default parameters was used. All attention multiple instance learning models were trained with a batch size of one. The rest of the hyperparameters (epochs, learning rate, learning rate decay routine between cosine annealing or multi-step, weight decay, dropout rate) were finetuned for each task-encoder combination. Binary cross-entropy loss was used for the classification tasks, and the negative log likelihood loss for the prognostic task.

### Statistical analysis

To analyze relationships between the feature extractor model performance and some potential drivers of this performance, we computed the correlation between the averaged performance across all tasks and the following variables: size of the model expressed in millions of parameters, number of training tiles, number of training WSIs, and number of training EC WSIs. Regarding the last variable, the authors of CONCH, H-OPTIMUS-0, and HIBOU-L did not report the number of EC WSIs or patients used in training. Hence, these models were excluded from this analysis. Given the likely non-linear nature of the relationships between these variables and the limited number of datapoints, we used the Spearman rank correlation instead of the Pearson correlation.

### Packages

For each foundation model, we followed the authors’ instructions with regard to which Python and PyTorch versions were used to extract features. To train the downstream supervised tasks, Python 3.8 and Pytorch v.1.10.0 were used. Statistical tests were implemented with Scipy Python package (v.1.5.2), visualizations with Altair Python package (v.4.2.0), AUC with Scikit-learn Python package (v.1.0.2), and C-index with Scikit-survival (v.0.17.2). Pandas Python package (v.1.2.2) was used for the data split.

## Results

### Study design

We evaluated seven publicly available pan-cancer foundation models (H-OPTIMUS-0, GIGAPATH, VIRCHOW2, HIBOU-L, UNI, CONCH, and CTransPath) and two versions of an EC-specific feature extractor (EsVIT-1 and EsVIT-8) across eight supervised prediction tasks in EC, incorporating morphological, molecular and prognostic tasks, using the largest EC WSI database to date (Figure 1a). Given a bag of tile-level features, an attention multiple instance learning deep learning model is trained with a five-fold cross-validation routine for each EC task.

**Figure 1:**
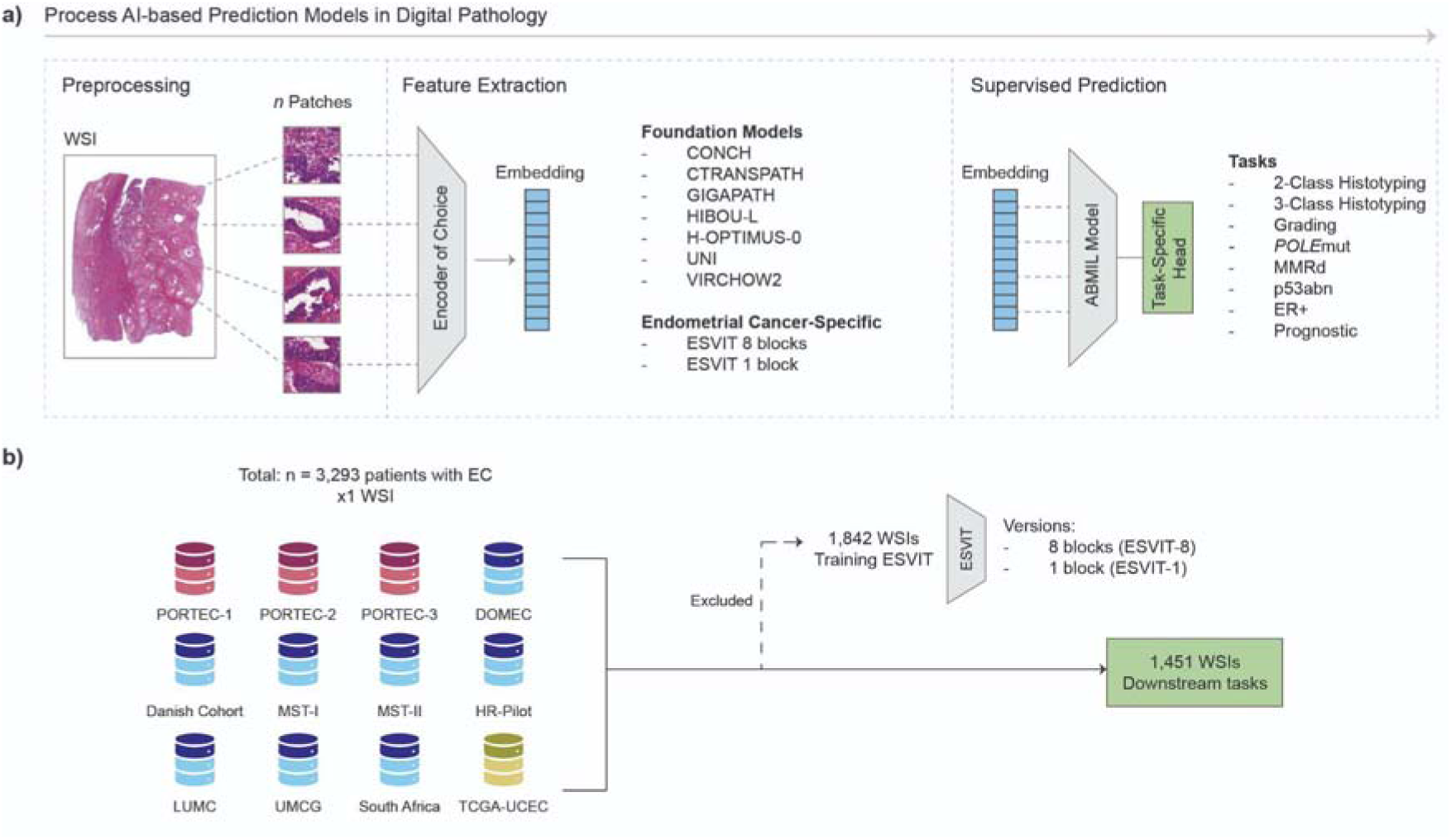
Overall representation of the study. **a)** Deep learning pipeline describing the experiments conducted in this study. First, digitized H&E-stained tumor slides of EC were tiled into fixed-size image tiles. Second, tile-level features are extracted using the pan-cancer foundation or EC-specific feature extractor models, yielding features with a model-dependent size described in Table 1. Third, tasks are assessed in this study that are important for clinical management in EC. Given a bag of tile-level features, an attention multiple instance learning deep learning model is trained with a five-fold cross-validation routine for each EC task. **b)** Collection of data: cohorts and division of WSIs for EC-specific feature extractor training and downstream tasks assessed in this study. Red: trial data, blue: clinical cohorts, green: public data.

**Table 1.**
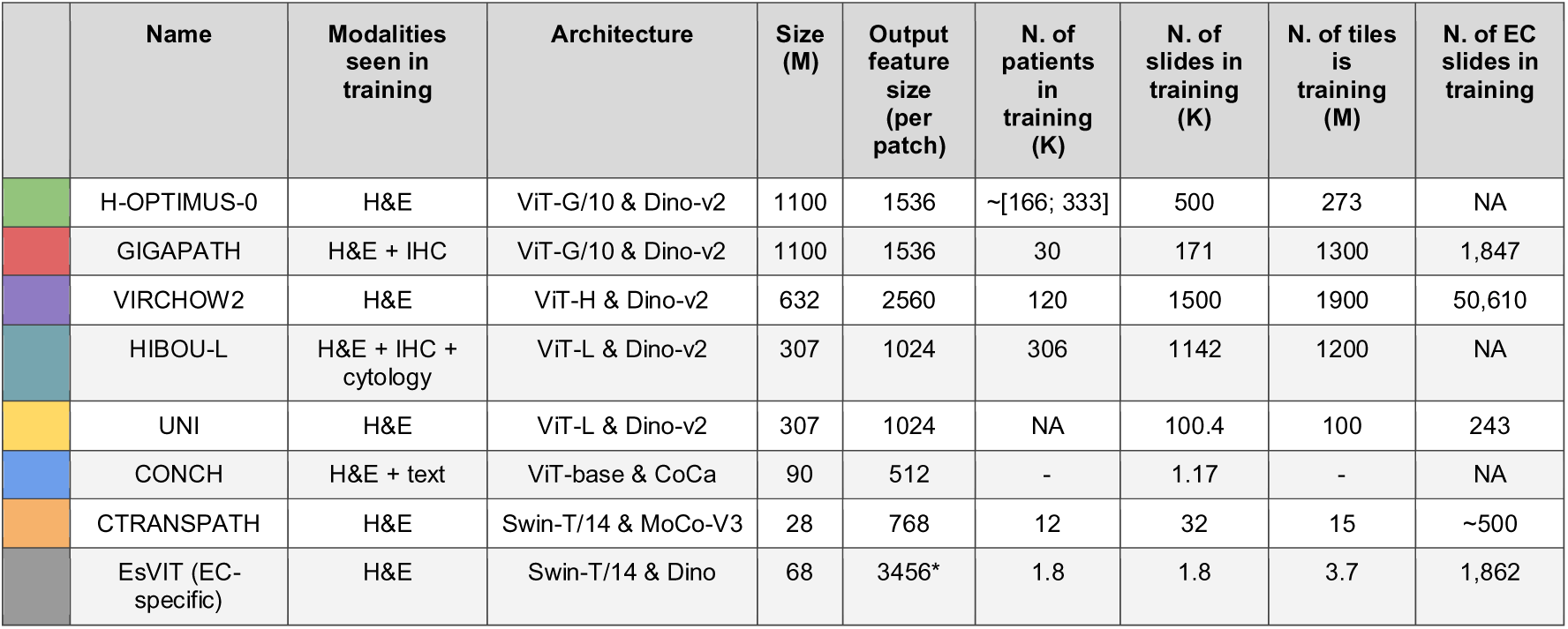
Characteristics of each feature extractor model used in this study. Information not disclosed in the publication is denoted with NA. The number of patients and slides used in training is reported in thousands (K), the number of training tiles in millions (M), and the model size in millions of parameters. For CONCH, an image-to-caption pair model, we considered the dataset of images used to train CONCH as slide images. H&E: hematoxylin and eosin, IHC: immunohistochemistry, ViT: vision transformers (G: giant, H: huge, L: large), Dino: distillation with no labels, CoCa: contrastive captioners, Swin: Swin transformer (T: tiny), MoCo: momentum contrast. *The final 8 transformer blocks were concatenated.

|  | Name | Modalities seen in training | Architecture | Size (M) | Output feature size (per patch) | N. of patients in training (K) | N. of slides in training (K) | N. of tiles in training (M) | N. of EC slides in training |
| --- | --- | --- | --- | --- | --- | --- | --- | --- | --- |
|  | H-OPTIMUS-0 | H&E | ViT-G/10 & Dino-v2 | 1100 | 1536 | ~[166; 333] | 500 | 273 | NA |
|  | GIGAPATH | H&E + IHC | ViT-G/10 & Dino-v2 | 1100 | 1536 | 30 | 171 | 1300 | 1,847 |
|  | VIRCHOW2 | H&E | ViT-H & Dino-v2 | 632 | 2560 | 120 | 1500 | 1900 | 50,610 |
|  | HIBOU-L | H&E + IHC + cytology | ViT-L & Dino-v2 | 307 | 1024 | 306 | 1142 | 1200 | NA |
|  | UNI | H&E | ViT-L & Dino-v2 | 307 | 1024 | NA | 100.4 | 100 | 243 |
|  | CONCH | H&E + text | ViT-base & CoCa | 90 | 512 | - | 1.17 | - | NA |
|  | CTRANSPATH | H&E | Swin-T/14 & MoCo-V3 | 28 | 768 | 12 | 32 | 15 | ~500 |
|  | EsViT (EC-specific) | H&E | Swin-T/14 & Dino | 68 | 3456* | 1.8 | 1.8 | 3.7 | 1,862 |

### Cohorts

Patients with EC were included from the three randomized trials (PORTEC-1^36^, PORTEC-2^37^, PORTEC-3^38^), one phase II trial (DOMEC^39^), seven clinical cohorts (Danish^40^, Medisch Spectrum Twente (MST)-I^41^, MST-II, transPORTEC high-risk pilot study^43^ (HR-Pilot), Leiden University Medical Center (LUMC)^18^, University Medical Center Groningen (UMCG)^44^, South-Africa^45^, and one publicly available dataset from the Cancer Genome Atlas Uterine Corpus (TCGA-UCEC)^46^ (Figure 1b). In total, 3,293 patients with EC were included, with one representative H&E-stained tumor-containing WSI each. Of the total, 1,842 WSIs (56%) were used for the training of the EC-specific feature encoder (EsVIT) in a previous study.^18,49^ The remaining 1,451 WSIs (54%) were used for the downstream tasks, the specific number depending on the task and the available ground truth data. No patient or WSI overlap existed between feature extractor training datasets and downstream task evaluation. Ground-truth labels were derived conforming to diagnostic workflows, and patients without available ground-truth data were excluded at the task level (Methods; Tables S2–S12).

### Task definitions

Morphological tasks comprised tumor grading (low-grade vs. high-grade; n = 1,036), two-class histological subtyping (endometrioid vs. non-endometrioid EC; n = 1,436), and three-class subtyping (endometrioid, serous, and clear cell carcinoma; n = 1,310). Molecular tasks included prediction of *POLE* mutation (*POLE*mut) status (n = 1,026), mismatch repair deficiency (MMRd; n = 1,137), abnormal p53 expression (p53abn; n = 1,137), and estrogen receptor status (ER+ vs. ER−). The specific numbers per class are depicted in Tables S2-S3, and per cohort in Tables S5-S11. The prognostic task consisted of predicting distant recurrence-free survival in stage I–III EC patients who did not receive adjuvant systemic therapy (n = 849; six-year median follow-up; 122 recurrences), using a discrete-time survival modeling approach (detailed composition per class in Table S4 and per cohort in Table S12). Evaluation metrics included area-under-the-curve (AUC) for classification tasks and the C-index for the prognostic task using the mean on the 5 cross-validation folds.

### Feature encoders

The foundation models were selected based on reported high performance (H-OPTIMUS-0, GIGAPATH, VIRCHOW2, UNI, CONCH), earlier development (CTransPath), or more recent release (HIBOU-L). The EC-specific feature extractor EsVIT was previously trained on 3.7 million EC tiles from 1,842 WSIs. We define EsVIT-1 as the version where the final transformer block was extracted as output features, and EsVIT-8 where the final 8 transformer blocks were concatenated and extracted as output features. The evaluated feature extractors differ in model scale (up to 1.1 billion parameters), training dataset size, and training modality, including image-text contrastive learning for CONCH (Table 1).

### Performance overall and across morphological, molecular and prognostic categories

Across all tasks, the evaluated foundation models showed consistently strong performance, with several achieving an area under the ROC curve (AUC) > 0.800. Among them, H-OPTIMUS-0, CONCH, and VIRCHOW2 ranked as the top-performing models overall, with mean metrics of 0.864 ± 0.073, 0.862 ± 0.066, and 0.860 ± 0.072, respectively (Table S1). The overall weakest performing foundation models were CTRANSPATH and HIBOU-L (mean metrics = 0.817 ± 0.095 and 0.821 ± 0.095, respectively), each ranking lowest in four out of eight tasks. Evaluating robustness using the variability of the performance across the cross-validation folds, UNI was the most stable in performance (standard deviation in AUC of 0.062), whereas HIBOU-L was the least stable (standard deviation in AUC of 0.095). By task category, we found that CONCH performed best for the morphological tasks with an average AUC of 0.910 ± 0.024, H-OPTIMUS-0 performed best for molecular tasks (*POLE*mut, p53abn, top-two for MMRd; average AUC of 0.875 ± 0.035), and UNI achieved the highest performance for the prognostic task (C-index = 0.727 ± 0.069) (Table S2-S4, Figure 2a).

**Figure 2:**
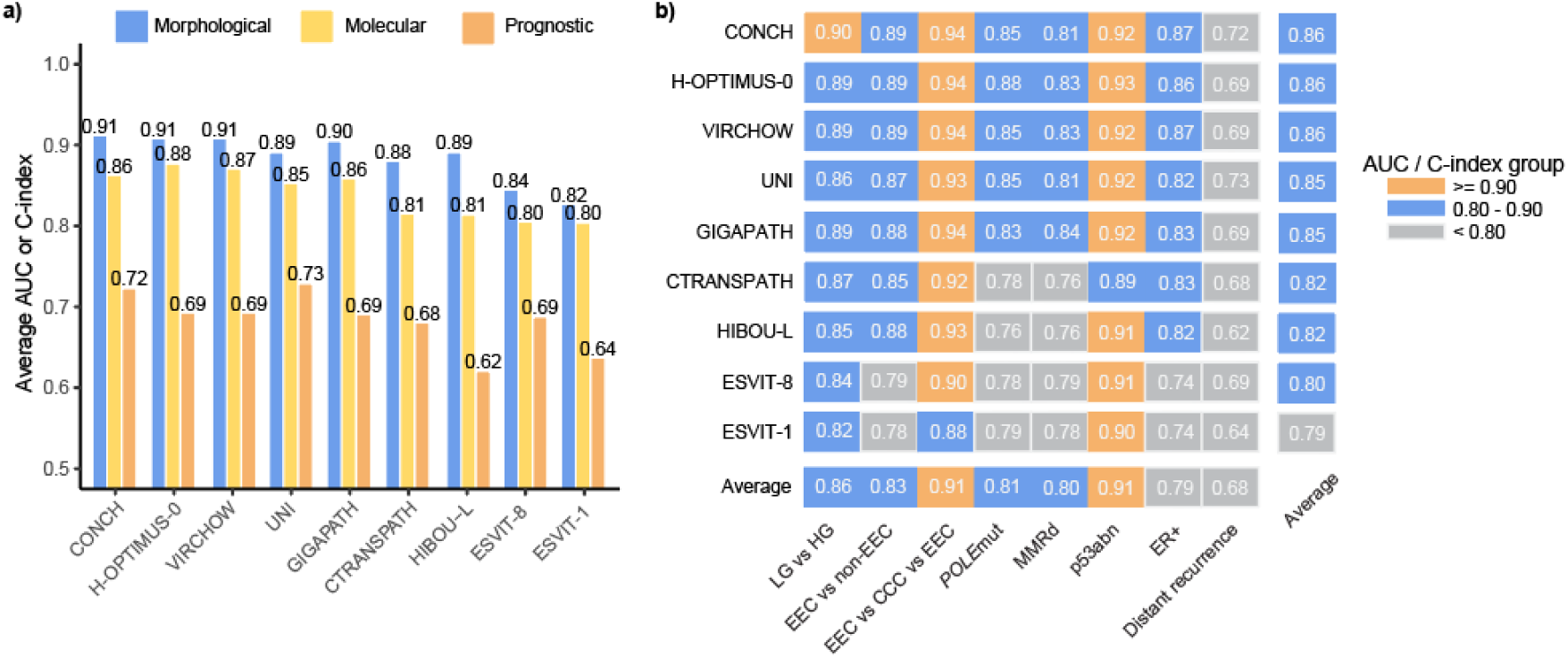
Performance of all feature extractors across morphologic, molecular and prognostic tasks. Exact numbers are reported in Table S1-S3, barplots per task are depicted in Figure S1. Note: for the distance recurrence task, the performance metric is the C-index.

The EC-specific encoders performed worse than the foundation models averaged across all tasks (average EsVIT: AUC = 0.834 ± 0.009; average foundation models: AUC = 0.897 ± 0.026). Averaged on the morphological and molecular tasks, they underperformed compared to the weakest foundation model. Only in the prognostic task, EsVIT-8 showed similar performances (AUC = 0.686 ± 0.131).

We granularized model performances per task based on AUC (or C-index for prognosis) as very high (>0.90), high (0.80–0.90), or low-intermediate (<0.80) (Figure 2b). Across morphological tasks, CONCH consistently reached high-performance levels in grading and three-class subtyping. Across molecular tasks, CONCH, H-OPTIMUS-0, VIRCHOW, UNI, GIGAPATH achieved at least the high AUC group in the tasks, and very high performance in p53abn prediction. All feature encoders achieved low-intermediate performance (AUC < 0.80) in the prognostic task. Weaker foundation models, including CTRANSPATH and HIBOU-L, frequently fell into low-intermediate categories (prediction of *POLE*mut, MMRd, distant recurrence). EsVIT encoders mostly achieved low-intermediate levels, with occasional very high performance in select molecular tasks (3-class histotyping, p53abn prediction).

### Performance of the feature encoders at task-level

For the morphological tasks, CONCH achieved the best performance for both grading (AUC = 0.900 ± 0.030) and three-class histologic subtyping (AUC = 0.944 ± 0.031) (Figure 3a, 3c; Table S2). VIRCHOW2 achieved top performance for the two-class histologic subtyping (EEC vs non-EEC, AUC = 0.892 ± 0.036) (Figure 3b). For the molecular tasks, H-OPTIMUS-0 performed best in *POLE*mut prediction (AUC = 0.876 ± 0.039) (Figure 3d; Table S3). This was a particularly notable advantage, exceeding the second-best model (VIRCHOW2) by a difference in AUC of 0.025. For *POLE*mut prediction, we found that the performance gap (Δ) between the least and best performing models was large (ΔAUC = 0.115) compared to other tasks. The MMRd classification task showed relatively small variation among the strongest models GIGAPATH, H-OPTIMUS-0, and VIRCHOW2 (AUCs = 0.839 ± 0.020, 0.835 ± 0.030, and 0.832 ± 0.047, respectively) (Figure 3e). The p53abn task showed minimal inter-model variability (standard deviation = 0.014), with H-OPTIMUS-0 performing the best (AUC = 0.930 ± 0.011) (Figure 3f). VIRCHOW2 achieved top performance in ER+ prediction (AUC = 0.868 ± 0.061) (Figure 3g). In the prognostic task, UNI achieved the highest performance (C-index = 0.727 ± 0.069) (Figure 3h; Table S4).

**Figure 3.**
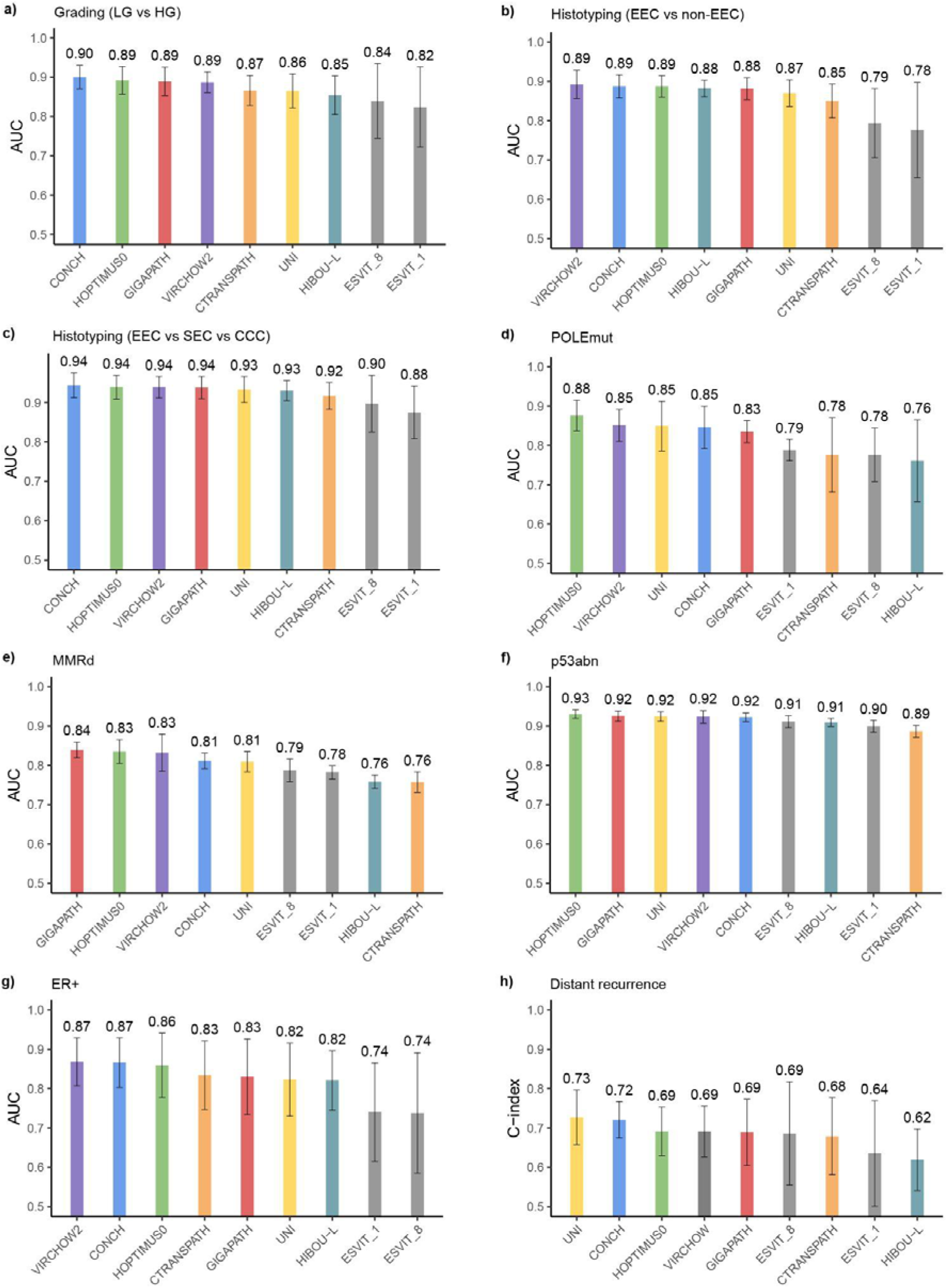
Barplot of performance of feature encoders per task. Mean values and error bars for standard deviation across folds are shown.

The two versions of EsVIT, the EC-specific feature encoders, were consistently outperformed by at least four foundation models on every task (Figure 3, Table S2-S4). In the morphological tasks, EsVIT performed worse than all foundation models, and showed large variability in performance across folds in cross-validation. Furthermore, in the molecular tasks, EsVIT underperformed compared to the weakest foundation model in ER+ (ΔAUC = −0.081) but performed slightly better than the weakest foundation model in the remaining molecular tasks (*POLE*mut ΔAUC = +0.027, MMRd ΔAUC = +0.030, p53abn ΔAUC = +0.025). EsVIT performed slightly better than the weakest foundation model in the prognostic task (ΔC-index = +0.067).

### Impact of model aspects on performance

To identify factors underlying the differences in performance identified in this study, we explored the correlations between each model’s average performance and key aspects: model size, training dataset size (tiles, WSIs), and number of EC WSIs (Figure 4). Model performance correlated positively with model size (Spearman IZl = 0.71, *P* = 0.032) and with the number of WSIs (IZl = 0.73, *P* = 0.026) and tiles (IZl = 0.73, *P* = 0.040) used for training. Notably, CONCH achieved high discriminative ability despite a comparatively smaller patch count. No meaningful correlation with performance was found for the number of EC slides included in the training data (IZl = 0.06, *P* = 0.913).

**Figure 4:**
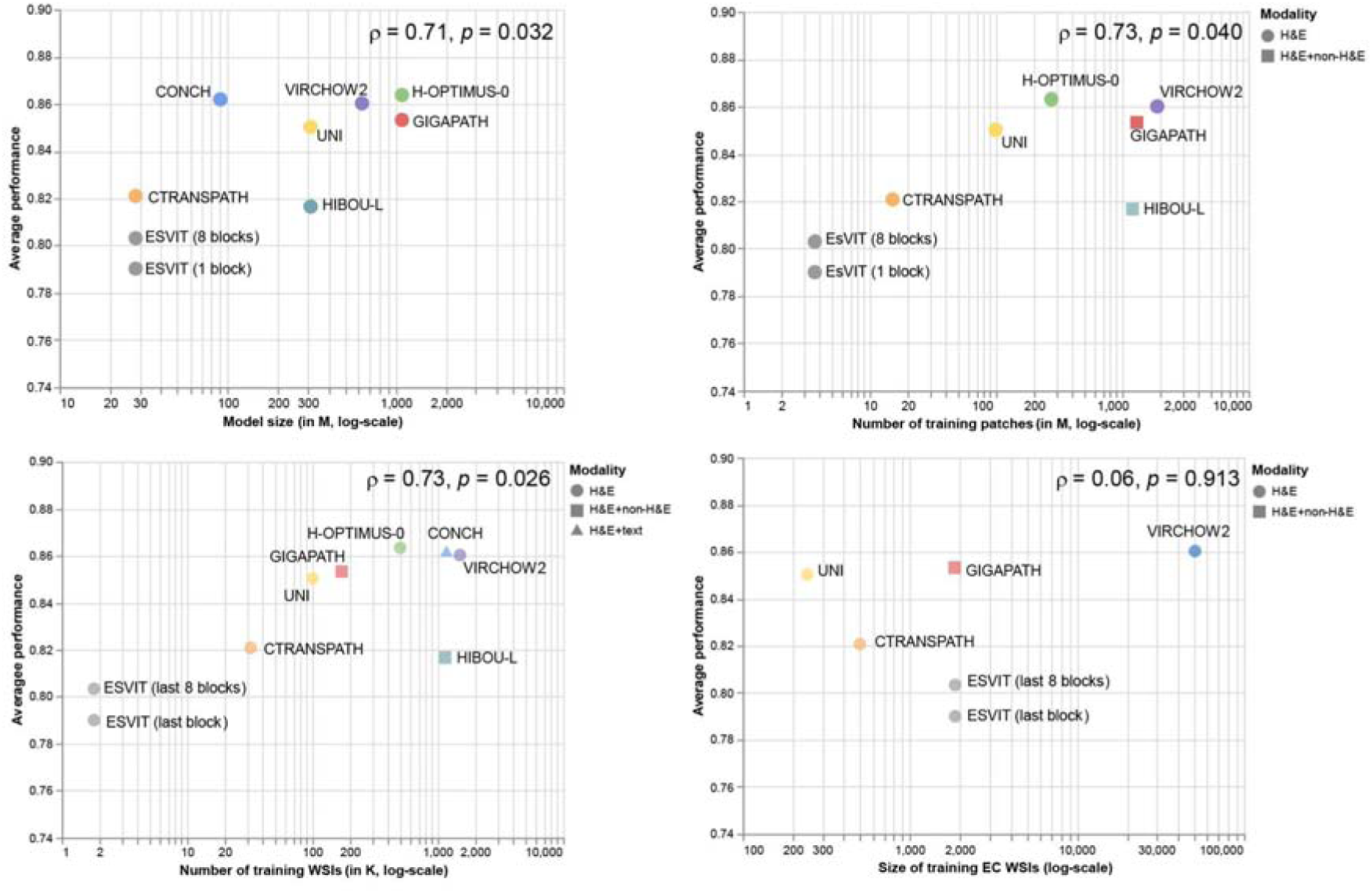
Impact of model aspects on performance. The comparison of the average performance of each feature extractor model against the model size (in millions of parameters (top-left)), and the size of the training dataset (in millions of tiles (top-right); in thousands of WSIs (bottom left); in absolute number of EC WSIS (bottom right)). The x-axis is shown on a log scale. The y-axis shows the averaged performance (mean) across all tasks. The number of EC WSIs included in the training dataset was not reported for CONCH, HIBOU-L, and H-OPTIMUS-0.

## Discussion

In this study, we leverage the largest EC cohort analyzed to date, to perform the first systematic comparison of state-of-the-art foundation models across eight clinically-relevant EC applications. We further benchmarked these models against an EC-specific feature extractor trained exclusively on EC WSIs. Overall, our results indicate that foundation models achieve performance levels that are potentially suitable for clinical use for routine H&E-based diagnostic tasks such as histological grading and subtyping. They demonstrate encouraging yet more variable performance for selected molecular prediction tasks, notably the p53abn prediction task reaching the same performance level as the morphological tasks. In contrast, prognostic modeling based solely on WSI-derived features remained substantially more challenging even with the use of foundation models, suggesting that outcome prediction tasks likely require multimodal integration of genomic, clinical, and imaging data as demonstrated by Volinsky-Fremond et al.18. Taken together, these findings underscore both the strength and the current limitations of foundation model-based representations.

Among the tested foundation models, H-OPTIMUS-0, CONCH, and VIRCHOW2 achieved the highest overall performance, which is in line with other benchmarking studies^26,29,32,34,51,52^. No single model consistently dominated across all tasks. Differences among the top models were marginal (approximately 1%), suggesting that additional training data may not always yield improvements in domain specific tasks. Notably, CONCH, a vision-text model^26^, achieved comparable results to the leading models despite being trained on fewer curated images. This finding suggests that integrating diverse data modalities may enhance generalization more effectively than further scaling dataset size alone, as proven by the success of multimodal foundation models such as MUSK^33^, THREADS^53^, and PRISM^54^.

Interestingly, the degree of performance variability across foundation models differed between tasks. For some tasks, such as grading, 3-class histotyping and p53abn prediction, performances were consistently above a 0.85 AUC across models, potentially suggesting that morphological correlates of these targets are captured. For others, like *POLE*mut prediction, larger differences were observed, which may indicate that only certain encoders effectively learned the relevant histomorphological patterns. Further analysis is required to confirm these hypotheses.

When compared to the EC-specific encoder, all foundation models showed superior performance. While tissue-specific models could outperform foundation models^52^, we found that the EC-specific extractor surpassed only the worst-performing foundation models in a few EC tasks, and it was consistently outperformed by the top-performing models across all tasks. The EC-specific model (EsVIT) demonstrated that disease-specific pretraining can capture relevant EC morphologies. Yet, foundation models’ pan-cancer representations achieved stronger generalization. Further work is needed to understand how data scale, diversity, and model capacity affect the generalizability of disease-specific encoders.

We investigated scaling laws by exploring the relationship between model performance, parameter size, and training data composition and size. We observed that overall performance correlated positively with both model size and training dataset size. This is in contrast to Campanella et al.^52^, who found that model size has a limited effect on model performance and that training data size did not affect model performance. Similarly, Neidlinger et al.^51^ found that larger training sets did not always yield better results. Campanella and Neidlinger both emphasized that the diversity of training samples may be a more critical determinant of model generalization. By investigating the relationship between performance and the EC WSI contribution to pretraining, we found that the number of EC slides used during pretraining did not affect model performance, suggesting that general morphological diversity, rather than disease-specific sample size, drives model generalizability in EC.

Methodological choices, including patch size, resolution, and aggregation strategy, were standardized to ensure fair comparison across models. While these choices may influence absolute performance, consistent settings allow meaningful relative comparisons. Future studies could explore alternative pre-processing settings, as well as cross-modality generalization of foundation models to immunohistochemistry, cytology, or other histopathological domains. Further investigation into model explainability, particularly the representations learned by the encoders, alongside robustness to input distribution shifts and rigorous external validation across diverse cohorts, remains essential for clinical translation.

In conclusion, publicly available foundation models represent a valuable resource for future AI diagnostic tool development in EC. We show that foundation models outperform EC-specific feature extractors across a broad range of clinically relevant prediction tasks while requiring minimal additional resources. While no single foundation model achieves the best performance for all tasks, our results provide guidance for selecting top-performing models based on the diagnostic or prognostic task. Although we cannot assume that these specific performance outcomes will directly translate to other cancers, the study design can serve as a framework for similar assessments in other disease domains. This would allow the community to accelerate clinically useful model development across histopathology. Collectively, these findings underscore the potential of foundation models to advance computational pathology and support precision diagnostics within EC and beyond.

### Data statement

The tumor material and datasets generated during or analyzed in the present study are not publicly available owing to restrictions in patient consent and privacy laws. Data and tumor material from PORTEC-1 and −2 are held by the PORTEC study group. PORTEC-3 and the TransPORTEC study is held by the international TransPORTEC consortium. MST-I/II and DOMEC are held by the LUMC researchers (C.C.R, T.B, N.H, J.R.K.). Data and tumor material from the Danish cohort are held by the coauthor of this article, G.Ø. Data and tumor material from the UMCG cohort are held by the coauthors of this article, H.W.N. Data and tumor material from the South-Africa cohort are held by the coauthor of this article R.R.R. Requests for sharing of all data should be addressed to the corresponding author within 15 years of the date of publication of this article and include a scientific proposal. Depending on the specific research proposal, the TransPORTEC consortium (PORTEC-3 and TransPORTEC study) or the PORTEC study group (PORTEC-1, PORTEC-2) or coauthors C.C.R, G.Ø., H.W.N., or N.H. and T.B., will determine when, for how long, for which specific purposes and under which conditions the requested data can be made available, subject to ethical consent. TCGA-UCEC images, mutational status and clinical data are publicly available via the cBioPortal for Cancer Genomics at https://www.cbioportal.org/study/clinicalData?id=ucec_tcga_pan_can_atlas_2018.

## Contributions

S.V.-F., N.H., V.H.K. and T.B. conceived the study design. S.V.-F. and N.vdB trained the models. J.B.W. provided technical support. S.V.-F, N.vdB, C.L.C., and, N.H. acquired the data. S.V.-F. analyzed the data. S.V.-F. and N.vdB drafted the paper and the figures. S.V.-F., N.vdB, J.BW, L.A.S, S.A, V.H.K., N.H, and T.B. substantially reviewed the paper. All authors critically reviewed the paper and the results and approved the final version.

## Competing interests statement

Authors declare no competing interests related to this study.

## Data Availability

The tumor material and datasets generated during or analyzed in the present study are not publicly available owing to restrictions in patient consent and privacy laws. Data and tumor material from PORTEC-1 and −2 are held by the PORTEC study group. PORTEC-3 and the TransPORTEC study is held by the international TransPORTEC consortium. MST-I/II and DOMEC are held by the LUMC researchers (C.C.R, T.B, N.H, J.R.K.). Data and tumor material from the Danish cohort are held by the coauthor of this article, G.O. Data and tumor material from the UMCG cohort are held by the coauthors of this article, H.W.N. Data and tumor material from the South-Africa cohort are held by the coauthor of this article R.R.R. Requests for sharing of all data should be addressed to the corresponding author within 15 years of the date of publication of this article and include a scientific proposal. Depending on the specific research proposal, the TransPORTEC consortium (PORTEC-3 and TransPORTEC study) or the PORTEC study group (PORTEC-1, PORTEC-2) or coauthors C.C.R, G.O., H.W.N., or N.H. and T.B., will determine when, for how long, for which specific purposes and under which conditions the requested data can be made available, subject to ethical consent. TCGA-UCEC images, mutational status and clinical data are publicly available via the cBioPortal for Cancer Genomics.

https://www.cbioportal.org/study/clinicalData?id=ucec_tcga_pan_can_atlas_2018.

## Acknowledgements

We are grateful to all patients, investigators, and participating hospitals, as well as the consortia and researchers who curated and shared clinical cohorts contributing to this study. We thank Diantha Terlouw, Leiden University Medical Center, for assistance with interpretation of NGS results, and Natalja ter Haar and Tessa Rutte, Leiden University Medical Center, for excellent technical support. We acknowledge the AIRMEC Consortium for the exceptional collaboration. The PORTEC-1, PORTEC-2, and PORTEC-3 trials were supported by grants from the Dutch Cancer Society (CKTO 90-01, CKTO 2001-04, CKTO 2006-04, respectively). We acknowledge the PORTEC-3 study group and the TransPORTEC Research Consortium for establishing and maintaining the TransPORTEC biobank. We also acknowledge the Netherlands Cancer Institute for access to their 3D-HISTECH P1000 scanner. We thank Lisa Vermij, Alicia León-Castillo and Ellen Stelloo for the contribution to molecularly classifying the samples. We acknowledge Famke Wakkerman for retrieving additional data for the LUMC cohort and maintaining the collected database. We acknowledge the SHARK team, the computational cluster of the LUMC, for their technical support. This work was supported by The Hanarth Foundation (no specific grant number).

## Notes

### Competing Interest Statement

The authors have declared no competing interest.

### Author Declarations

The PORTEC1/2/3 study protocols were approved by the Medical Ethical Committee Leiden (Den Haag) Delft and the medical ethics committees at participating centers. Both studies were conducted in accordance with the principles of the Declaration of Helsinki. All patients provided signed informed consent to study participation. For the retrospective use of the clinical trials and retrospective cohorts (TransPORTEC study, MST), we have obtained ethical permission to use the data by the Medical Ethical Committee Leiden (Den Haag) Delft (B21.065 and B21.011) as well as the Leiden Cohort (nWMO‐D4‐2023‐002), and the Danish Cohort by the Center for Regional Udvikling De Videnskabsetiske Komiteer(H16025909).

