## Supplementary material for "Assessing Foundation Models for Computational Pathology in Endometrial Cancer"

### Supplementary Information

**Performance overview** **2**
All tasks 2
Morphological tasks 3
Molecular tasks 4
Prognostic task 5
**Cohort overview** **6**
Endometrioid versus Non-Endometrioid task 6
Endometrioid versus Serous versus Clear Cell task 7
Low-grade (grade 1-2) Endometrioid versus High-grade (grade 3) Endometrioid task 8
*POLE*mut versus wildtype task 9
MMRd versus MMRp task 10
p53abn versus wildtype task 11
ER positive versus negative task 12
Distant recurrence task 13

### Performance overview

#### All tasks

##### Table S1 Average performance across all tasks.

| | Task | Average performance (all tasks) |
|---|---|---|
| | HOPTIMUS0 | 0.864 ± 0.073 |
| | CONCH | 0.862 ± 0.066 |
| | VIRCHOW | 0.860 ± 0.072 |
| | GIGAPATH | 0.853 ± 0.073 |
| | UNI | 0.850 ± 0.062 |
| | CTRANSPATH | 0.821 ± 0.073 |
| | HIBOU-L | 0.817 ± 0.095 |
| | EsVIT (concatenation last 8 blocks) | 0.803 ± 0.071 |
| | EsVIT (final block) | 0.790 ± 0.077 |

#### Morphological tasks

**Table S2: Performance of the seven foundation models and two EC-specific feature extractors on H&E-based morphological tasks.** Performance is reported as mean and standard deviation across five-fold cross-validation and computed with the AUC. EC = Endometrial cancer; LG = Low grade; HG = High grade; EEC = Endometrioid; SEC = Serous; CCC = Clear Cells.

| | Task | LG EEC vs HG EEC | EEC vs non-EEC | EEC vs SEC vs CCC | Average per model |
|---|---|---|---|---|---|
| | N. of patients (%) | Total 1,036<br>- LG: 736 (71%)<br>- HG: 300 (29%) | Total 1,436<br>- EEC: 1,036 (72%)<br>- non-EEC: 400 (28%) | Total 1,310<br>- EEC: 1,036 (79%)<br>- SEC: 202 (15%)<br>- CCC: 72 (5%) | |
| | CONCH | 0.900 ± 0.030 | 0.887 ± 0.029 | 0.944 ± 0.031 | 0.910 ± 0.024 |
| | H-OPTIMUS-0 | 0.892 ± 0.035 | 0.887 ± 0.027 | 0.939 ± 0.030 | 0.906 ± 0.023 |
| | VIRCHOW2 | 0.887 ± 0.026 | 0.892 ± 0.036 | 0.939 ± 0.027 | 0.906 ± 0.023 |
| | GIGAPATH | 0.889 ± 0.036 | 0.881 ± 0.028 | 0.938 ± 0.028 | 0.903 ± 0.025 |
| | UNI | 0.865 ± 0.043 | 0.870 ± 0.034 | 0.933 ± 0.033 | 0.889 ± 0.031 |
| | HIBOU-L | 0.854 ± 0.049 | 0.882 ± 0.021 | 0.930 ± 0.026 | 0.889 ± 0.031 |
| | CTRANSPATH | 0.866 ± 0.038 | 0.850 ± 0.043 | 0.917 ± 0.034 | 0.878 ± 0.029 |
| | **Average foundation models per task** | 0.879 ± 0.016 | 0.878 ± 0.013 | 0.934 ± 0.008 | 0.897 ± 0.026 |
| | EsVIT (concatenation last 8 blocks) | 0.839 ± 0.095 | 0.793 ± 0.088 | 0.897 ± 0.072 | 0.843 ± 0.043 |
| | EsVIT (final block) | 0.824 ± 0.102 | 0.776 ± 0.121 | 0.875 ± 0.067 | 0.825 ± 0.040 |
| | **Average EC-specific models per task** | 0.832 ± 0.008 | 0.785 ± 0.009 | 0.886 ± 0.011 | 0.834 ± 0.009 |

#### Molecular tasks

**Table S3: Performance of the seven foundation models and two EC-specific feature extractors on molecular tasks.** Performance is reported as mean and standard deviation across five-fold cross-validation and computed with the AUC. EC = Endometrial cancer; *POLE*mut = *POLE* mutant; MMRd = Mismatch repair deficient; MMRp = Mismatch repair proficient; p53abn = p53 abnormal; wt = wildtype; ER = Estrogen Receptor.

| | **Task** | ***POLE*mut vs *POLE*wt** | **MMRd vs MMRp** | **p53abn vs p53wt** | **ER+ vs ER-** | **Average per model** |
|---|---|---|---|---|---|---|
| | N. of patients (%) | Total 1,026<br>- *POLE*mut: 81 (8%)<br>- *POLE*wt: 924 (92%) | Total 1,137<br>- MMRd: 306 (27%)<br>- MMRp: 831 (73%) | Total 1,137<br>- p53abn: 265 (23%)<br>- p53wt: 872 (77%) | Total 1,152<br>- ER+: 884 (77%)<br>-ER-: 268 (23%) | |
| | H-OPTIMUS-0 | 0.876 ± 0.039 | 0.835 ± 0.030 | 0.930 ± 0.011 | 0.859 ± 0.082 | 0.875 ± 0.035 |
| | VIRCHOW2 | 0.851 ± 0.041 | 0.832 ± 0.047 | 0.923 ± 0.016 | 0.868 ± 0.061 | 0.869 ± 0.034 |
| | CONCH | 0.846 ± 0.054 | 0.811 ± 0.020 | 0.922 ± 0.011 | 0.866 ± 0.063 | 0.861 ± 0.040 |
| | GIGAPATH | 0.835 ± 0.028 | 0.839 ± 0.020 | 0.925 ± 0.013 | 0.830 ± 0.096 | 0.857 ± 0.039 |
| | UNI | 0.849 ± 0.063 | 0.809 ± 0.026 | 0.924 ± 0.012 | 0.823 ± 0.093 | 0.851 ± 0.044 |
| | CTRANSPATH | 0.776 ± 0.094 | 0.757 ± 0.026 | 0.886 ± 0.015 | 0.834 ± 0.087 | 0.813 ± 0.051 |
| | HIBOU-L | 0.761 ± 0.104 | 0.758 ± 0.017 | 0.909 ± 0.010 | 0.821 ± 0.076 | 0.812 ± 0.061 |
| | **Average foundation models per task** | 0.828 ± 0.039 | 0.806 ± 0.032 | 0.917 ± 0.014 | 0.843 ± 0.019 | 0.848 ± 0.024 |
| | EsVIT (concatenation last 8 blocks) | 0.776 ± 0.068 | 0.787 ± 0.029 | 0.911 ± 0.015 | 0.738 ± 0.153 | 0.803 ± 0.065 |
| | EsVIT (final block) | 0.788 ± 0.027 | 0.782 ± 0.017 | 0.899 ± 0.015 | 0.740 ± 0.125 | 0.802 ± 0.059 |
| | **Average EC-specific models per task** | 0.782 ± 0.006 | 0.785 ± 0.003 | 0.905 ± 0.006 | 0.739 ± 0.001 | 0.803 ± 0.000 |

#### Prognostic task

**Table S4: Performance of the seven foundation models and two EC-specific feature extractors on the prognostic task.** Performance is reported as mean and standard deviation across five-fold cross-validation and computed with the C-index. EC = Endometrial cancer.

| | Task | Distant recurrence |
|---|---|---|
| | N. of patients (%) | Total 849<br>- recurrence: 122 (14%)<br>- censored: 727 (86%) |
| | Median follow-up (years) | 6 |
| | UNI | 0.727 ± 0.069 |
| | CONCH | 0.721 ± 0.046 |
| | HOPTIMUS0 | 0.691 ± 0.062 |
| | VIRCHOW | 0.691 ± 0.065 |
| | GIGAPATH | 0.689 ± 0.084 |
| | CTRANSPATH | 0.679 ± 0.098 |
| | HIBOU | 0.619 ± 0.078 |
| | **Average foundation models per task** | 0.689 ± 0.033 |
| | EsVIT (concatenation last 8 blocks) | 0.686 ± 0.131 |
| | EsVIT (final block) | 0.635 ± 0.134 |
| | **Average EC-specific models per task** | 0.661 ± 0.021 |

#### Cohort overview

##### Endometrioid versus Non-Endometrioid task

| | inclusion | PORTEC-1 | PORTEC-2 | PORTEC-3 | transPORTEC | MST-I | MST-II | DOMEC | DANISH | LUMC | UMCG | SOUTH-AFRICA | Total | %of included |
|---|---|---|---|---|---|---|---|---|---|---|---|---|---|---|
| **Number of patients** | | | | | | | | | | | | | | |
| | yes | 120 | 100 | 268 | 28 | 52 | 172 | 45 | 142 | 197 | 172 | 140 | **1436** | - |
| | no | 594 | 327 | 392 | 88 | 219 | 24 | 10 | 270 | 25 | 106 | 10 | 2065 | - |
| **Age: median** | | | | | | | | | | | | | | |
| | yes | 67 | 69 | 62 | 66 | 64 | 68 | - | 69 | 68 | 65 | 65 | **67** | - |
| | no | 66 | 70 | 62 | 68 | 70 | 65 | - | 70 | 64 | 63 | 63 | **66** | - |
| **Histotype** | | | | | | | | | | | | | | |
| Endometrioid grade 1-2 | yes | 95 | 85 | 104 | 4 | 21 | 132 | 9 | 0 | 101 | 156 | 29 | **736** | **51.3** |
| | no | 506 | 289 | 153 | 14 | 80 | 0 | 1 | 0 | 12 | 0 | 1 | 1056 | - |
| Endometrioid grade 3 | yes | 19 | 9 | 73 | 20 | 16 | 22 | 5 | 64 | 28 | 8 | 36 | **300** | **20.9** |
| | no | 76 | 32 | 112 | 48 | 53 | 1 | 1 | 128 | 4 | 0 | 1 | 456 | - |
| Serous carcinoma | yes | 1 | 6 | 44 | 3 | 2 | 8 | 16 | 54 | 29 | 7 | 32 | **202** | **14.1** |
| | no | 8 | 5 | 61 | 9 | 23 | 0 | 5 | 91 | 6 | 0 | 2 | 210 | - |
| Clear cell carcinoma | yes | 2 | 0 | 27 | 1 | 2 | 2 | 6 | 13 | 15 | 0 | 4 | **72** | **5.0** |
| | no | 2 | 0 | 35 | 17 | 12 | 0 | 0 | 23 | 1 | 0 | 1 | 91 | - |
| Carcinosarcoma | yes | 0 | 0 | 0 | 0 | 3 | 3 | 6 | 4 | 11 | 0 | 33 | **60** | **4.2** |
| | no | 0 | 0 | 0 | 0 | 21 | 0 | 2 | 9 | 1 | 0 | 5 | 38 | - |
| Un-dedifferentiated | yes | 0 | 0 | 8 | 0 | 2 | 3 | 3 | 5 | 5 | 0 | 0 | **26** | **1.8** |
| | no | 0 | 0 | 6 | 0 | 7 | 0 | 0 | 12 | 1 | 0 | 0 | 26 | - |
| Other/Unknown | yes | 3 | 0 | 12 | 0 | 6 | 2 | 0 | 2 | 8 | 1 | 6 | **40** | **2.8** |
| | no | 2 | 1 | 25 | 0 | 23 | 23 | 1 | 7 | 0 | 106 | 0 | 188 | - |
| **LVSI** | | | | | | | | | | | | | | |
| Present | yes | 4 | 6 | 168 | 18 | 12 | 9 | 21 | 24 | 44 | 0 | 45 | **351** | **24.5** |
| | no | 22 | 14 | 221 | 37 | 41 | 0 | 1 | 31 | 4 | 0 | 0 | 371 | - |
| Focal or absent | yes | 102 | 90 | 100 | 10 | 39 | 159 | 21 | 118 | 137 | 0 | 89 | **865** | **60.4** |
| | no | 435 | 283 | 171 | 30 | 163 | 1 | 2 | 236 | 18 | 0 | 9 | 1348 | - |
| Unknown | yes | 14 | 4 | 0 | 0 | 1 | 4 | 3 | 0 | 16 | 172 | 6 | **220** | **15.1** |
| | no | 137 | 30 | 0 | 21 | 15 | 23 | 7 | 3 | 3 | 106 | 1 | 346 | - |
| **2009 FIGO stage** | | | | | | | | | | | | | | |
| IA | yes | 47 | 16 | 35 | 3 | 2 | 35 | 0 | 31 | 85 | 114* | 47 | **415** | **60.2*** |
| | no | 247 | 55 | 43 | 8 | 26 | 9 | 0 | 143 | 10 | 63* | 3 | 607 | - |
| IB | yes | 73 | 83 | 47 | 6 | 11 | 134 | 0 | 27 | 43 | - | 24 | **448** | - |
| | no | 347 | 268 | 70 | 25 | 63 | 15 | 0 | 69 | 6 | - | 1 | 864 | - |
| II | yes | 0 | 0 | 64 | 5 | 12 | 0 | 0 | 7 | 14 | 19 | 23 | **144** | **10.0** |
| | no | 0 | 2 | 106 | 16 | 70 | 0 | 0 | 22 | 0 | 13 | 1 | 230 | - |
| IIIA | yes | 0 | 0 | 32 | 5 | 16 | 0 | 0 | 4 | 14 | 33* | 10 | **114** | **22.4*** |
| | no | 0 | 2 | 51 | 13 | 36 | 0 | 0 | 2 | 0 | 23* | 3 | 130 | - |
| IIIB | yes | 0 | 1 | 15 | 0 | 8 | 0 | 0 | 5 | 3 | - | 1 | **33** | - |
| | no | 0 | 0 | 27 | 4 | 11 | 0 | 0 | 9 | 0 | - | 0 | 51 | - |
| IIIC | yes | 0 | 0 | 75 | 8 | 3 | 0 | 0 | 42 | 16 | - | 30 | **174** | - |
| | no | 0 | 0 | 95 | 11 | 12 | 0 | 0 | 19 | 1 | - | 0 | 138 | - |
| IV | yes | 0 | 0 | 0 | 1 | 0 | 0 | 45 | 26 | 2 | 6 | 0 | **80** | **5.6** |
| | no | 0 | 0 | 0 | 10 | 1 | 0 | 10 | 6 | 2 | 7 | 1 | 37 | - |
| Unkown | yes | 0 | 0 | 0 | 0 | 0 | 3 | 0 | 0 | 20 | 0 | 5 | **28** | **1.7** |
| | no | 0 | 0 | 0 | 1 | 0 | 0 | 0 | 0 | 6 | 0 | 1 | 8 | - |
| **Molecular class** | | | | | | | | | | | | | | |
| *POLE*mut | yes | 9 | 7 | 29 | 3 | 4 | 15 | 0 | 8 | 5 | 0 | 10 | **90** | **6.3** |
| | no | 33 | 17 | 22 | 13 | 12 | 0 | 0 | 30 | 0 | 0 | 0 | 127 | - |
| MMRd | yes | 22 | 20 | 77 | 6 | 10 | 54 | 7 | 37 | 48 | 0 | 36 | **317** | **22.1** |
| | no | 115 | 90 | 62 | 14 | 61 | 0 | 3 | 73 | 0 | 0 | 0 | 418 | - |
| p53abn | yes | 9 | 4 | 56 | 6 | 15 | 11 | 28 | 70 | 54 | 0 | 67 | **320** | **22.3** |
| | no | 31 | 26 | 43 | 28 | 54 | 1 | 6 | 120 | 1 | 0 | 6 | 316 | - |
| NSMP | yes | 52 | 64 | 74 | 13 | 21 | 80 | 9 | 27 | 77 | 0 | 24 | **441** | **30.8** |
| | no | 213 | 168 | 48 | 30 | 69 | 0 | 0 | 47 | 6 | 0 | 3 | 584 | - |
| Unkown | yes | 28 | 5 | 32 | 0 | 2 | 12 | 1 | 0 | 13 | 172 | 3 | **268** | **18.5** |
| | no | 202 | 26 | 217 | 3 | 23 | 23 | 1 | 0 | 18 | 106 | 1 | 620 | - |

**Table S5 Detailed composition by cohort of the supervised training dataset for the task Endometrioid versus Non-Endometrioid (2-class) histological subtype.** The inclusion variable represents the number of patients (with one WSI for each) included in the supervised training task. The percentage column is computed over the total number of included patients. For the UMCG cohort, only 2009 FIGO stage I, II, and III was known (as opposed to the subtype IA, IB, and IIIA, IIIB, IIIC), hence stage proportion numbers denoted with * represent pooled stage I or pooled stage III. *POLE*mut = *POLE* mutant; MMRd = Mismatch repair deficient; p53abn = p53 abnormal; NSMP = No specific molecular profile; LVSI = Lymphovascular space invasion.

#### Endometrioid versus Serous versus Clear Cell task

| | inclusion | PORTEC-1 | PORTEC-2 | PORTEC-3 | ansPORTE | MST-I | MST-II | DOMEC | DANISH | LUMC | UMCG | SOUTH-AFRICA | Total | % of included |
|---|---|---|---|---|---|---|---|---|---|---|---|---|---|---|
| **Number of patients** | | | | | | | | | | | | | | |
| | yes | 117 | 100 | 248 | 28 | 41 | 164 | 36 | 131 | 173 | 171 | 101 | **1310** | - |
| | no | 597 | 327 | 412 | 88 | 230 | 32 | 19 | 281 | 49 | 107 | 49 | 2191 | - |
| **Age: median** | | | | | | | | | | | | | | |
| | yes | 67 | 69 | 62 | 66 | 63 | 68 | - | 69 | 68 | 65 | 65 | **67** | - |
| | no | 66 | 70 | 62 | 68 | 70 | 65 | - | 70 | 66 | 63 | 67 | **67** | - |
| **Histotype** | | | | | | | | | | | | | | |
| Endometrioid grade 1-2 | yes | 95 | 85 | 104 | 4 | 21 | 132 | 9 | 0 | 101 | 156 | 29 | **736** | **56.2** |
| | no | 506 | 289 | 153 | 14 | 80 | 1 | 1 | 0 | 12 | 0 | 1 | 1057 | - |
| Endometrioid grade 3 | yes | 19 | 9 | 73 | 20 | 16 | 22 | 5 | 64 | 28 | 8 | 36 | **300** | **22.9** |
| | no | 76 | 32 | 112 | 48 | 53 | 1 | 1 | 128 | 4 | 0 | 1 | 456 | - |
| Serous carcinoma | yes | 1 | 6 | 44 | 3 | 2 | 8 | 16 | 54 | 29 | 7 | 32 | **202** | **15.4** |
| | no | 8 | 5 | 61 | 9 | 23 | 0 | 5 | 91 | 6 | 0 | 2 | 210 | - |
| Clear cell carcinoma | yes | 2 | 0 | 27 | 1 | 2 | 2 | 6 | 13 | 15 | 0 | 4 | **72** | **5.5** |
| | no | 2 | 0 | 35 | 17 | 12 | 0 | 0 | 23 | 1 | 0 | 1 | 91 | - |
| Carcinosarcoma | yes | 0 | 0 | 0 | 0 | 0 | 0 | 0 | 0 | 0 | 0 | 0 | **0** | **0.0** |
| | no | 0 | 0 | 0 | 0 | 24 | 3 | 8 | 13 | 12 | 0 | 38 | 98 | - |
| Un-dedifferentiated | yes | 0 | 0 | 0 | 0 | 0 | 0 | 0 | 0 | 0 | 0 | 0 | **0** | **0.0** |
| | no | 0 | 0 | 14 | 0 | 9 | 3 | 3 | 17 | 6 | 0 | 0 | 52 | - |
| Other/Unknown | yes | 0 | 0 | 0 | 0 | 0 | 0 | 0 | 0 | 0 | 0 | 0 | **0** | **0.0** |
| | no | 5 | 1 | 37 | 0 | 29 | 24 | 1 | 9 | 8 | 107 | 6 | 227 | - |
| **LVSI** | | | | | | | | | | | | | | |
| Present | yes | 4 | 6 | 149 | 18 | 8 | 9 | 15 | 20 | 35 | 0 | 36 | **300** | **23.0** |
| | no | 22 | 14 | 240 | 37 | 45 | 0 | 7 | 35 | 13 | 0 | 9 | 422 | - |
| Focal or absent | yes | 99 | 90 | 99 | 10 | 33 | 152 | 19 | 111 | 123 | 0 | 62 | **798** | **61.1** |
| | no | 438 | 283 | 172 | 30 | 169 | 9 | 4 | 243 | 32 | 0 | 36 | 1416 | - |
| Unknown | yes | 14 | 4 | 0 | 0 | 0 | 3 | 2 | 0 | 15 | 171 | 3 | **212** | **16.0** |
| | no | 137 | 30 | 0 | 21 | 16 | 23 | 8 | 3 | 4 | 107 | 4 | 353 | - |
| **2009 FIGO stage** | | | | | | | | | | | | | | |
| IA | yes | 45 | 16 | 30 | 3 | 2 | 28 | 0 | 29 | 81 | 114* | 33 | **381** | **61.7*** |
| | no | 249 | 55 | 48 | 8 | 26 | 16 | 0 | 145 | 14 | 63* | 17 | 641 | - |
| IB | yes | 72 | 83 | 39 | 6 | 8 | 133 | 0 | 25 | 39 | - | 21 | **426** | - |
| | no | 348 | 268 | 78 | 25 | 66 | 16 | 0 | 71 | 10 | - | 4 | 886 | - |
| II | yes | 0 | 0 | 62 | 5 | 11 | 0 | 0 | 7 | 12 | 19 | 17 | **152** | **11.6** |
| | no | 0 | 2 | 108 | 16 | 71 | 0 | 0 | 22 | 2 | 13 | 7 | 254 | - |
| IIIA | yes | 0 | 0 | 31 | 5 | 12 | 0 | 0 | 4 | 13 | 32* | 6 | **103** | **21.6*** |
| | no | 0 | 2 | 52 | 13 | 40 | 0 | 0 | 2 | 1 | 24* | 7 | 141 | - |
| IIIB | yes | 0 | 1 | 12 | 0 | 5 | 0 | 0 | 5 | 2 | - | 1 | **26** | - |
| | no | 0 | 0 | 30 | 4 | 14 | 0 | 0 | 9 | 1 | - | 0 | 58 | - |
| IIIC | yes | 0 | 0 | 74 | 8 | 3 | 0 | 0 | 36 | 13 | - | 19 | **153** | - |
| | no | 0 | 0 | 96 | 11 | 12 | 0 | 0 | 25 | 4 | - | 11 | 159 | - |
| IV | yes | 0 | 0 | 0 | 1 | 0 | 0 | 36 | 25 | 1 | 6 | 0 | **69** | **5.3** |
| | no | 0 | 0 | 0 | 10 | 1 | 0 | 19 | 7 | 3 | 7 | 1 | 48 | - |
| Unkown | yes | 0 | 0 | 0 | 0 | 0 | 3 | 0 | 0 | 12 | - | 4 | **19** | **1.2** |
| | no | 0 | 0 | 0 | 1 | 0 | 0 | 0 | 0 | 14 | - | 2 | 17 | - |
| **Molecular class** | | | | | | | | | | | | | | |
| *POLE* mut | yes | 9 | 7 | 27 | 3 | 2 | 14 | 0 | 8 | 5 | 0 | 9 | **84** | **6.4** |
| | no | 33 | 17 | 24 | 13 | 14 | 1 | 0 | 30 | 0 | 0 | 1 | 133 | - |
| MMRd | yes | 22 | 20 | 69 | 6 | 9 | 52 | 7 | 36 | 44 | 0 | 28 | **293** | **22.4** |
| | no | 115 | 90 | 70 | 14 | 62 | 2 | 3 | 74 | 4 | 0 | 8 | 442 | - |
| p53abn | yes | 8 | 4 | 49 | 6 | 10 | 8 | 20 | 62 | 37 | 0 | 39 | **243** | **18.6** |
| | no | 32 | 26 | 50 | 28 | 59 | 4 | 14 | 128 | 18 | 0 | 34 | 393 | - |
| NSMP | yes | 51 | 64 | 74 | 13 | 20 | 79 | 8 | 25 | 75 | 0 | 22 | **431** | **33.0** |
| | no | 214 | 168 | 48 | 30 | 70 | 1 | 1 | 49 | 8 | 0 | 5 | 594 | - |
| Unkown | yes | 27 | 5 | 29 | 0 | 0 | 11 | 1 | 0 | 12 | 171 | 3 | **259** | **19.6** |
| | no | 203 | 26 | 220 | 3 | 25 | 24 | 1 | 0 | 19 | 107 | 1 | 629 | - |

**Table S6 Detailed composition by cohort of the supervised training dataset for the task Endometrioid versus Serous versus Clear Cell (3-class) histological subtype.** The inclusion variable represents the number of patients (with one WSI for each) included in the supervised training task. The percentage column is computed over the total number of included patients. For the UMCG cohort, only 2009 FIGO stage I, II, and III was known (as opposed to the subtype IA, IB, and IIIA, IIIB, IIIC), hence stage proportion numbers denoted with * represent pooled stage I or pooled stage III. *POLE*mut = *POLE* mutant; MMRd = Mismatch repair deficient; p53abn = p53 abnormal; NSMP = No specific molecular profile; LVSI = Lymphovascular space invasion.

#### Low-grade (grade 1-2) Endometrioid versus High-grade (grade 3) Endometrioid task

| | inclusion | PORTEC-1 | PORTEC-2 | PORTEC-3 | transPORTEC | MST-I | MST-II | DOMEC | DANISH | LUMC | UMCG | SOUTH-AFRICA | Total | % of included |
|---|---|---|---|---|---|---|---|---|---|---|---|---|---|---|
| **Number of patients** | | | | | | | | | | | | | | |
| | yes | 114 | 94 | 177 | 24 | 37 | 154 | 14 | 64 | 129 | 164 | 65 | **1036** | - |
| | no | 600 | 333 | 483 | 92 | 234 | 42 | 41 | 348 | 93 | 114 | 85 | 2465 | - |
| **Age: median** | | | | | | | | | | | | | | |
| | yes | 67 | 69 | 60 | 64 | 63 | 68 | - | 68 | 64 | 65 | 59 | **65** | - |
| | no | 66 | 70 | 63 | 68 | 70 | 65 | - | 70 | 69 | 63 | 67 | **68** | - |
| **Histotype** | | | | | | | | | | | | | | |
| Endometrioid grade 1-2 | yes | 95 | 85 | 104 | 4 | 21 | 132 | 9 | 0 | 101 | 156 | 29 | **736** | **71.0** |
| | no | 506 | 289 | 153 | 14 | 80 | 1 | 1 | 0 | 12 | 0 | 1 | 1057 | - |
| Endometrioid grade 3 | yes | 19 | 9 | 73 | 20 | 16 | 22 | 5 | 64 | 28 | 8 | 36 | **300** | **29.0** |
| | no | 76 | 32 | 112 | 48 | 53 | 1 | 1 | 128 | 4 | 0 | 1 | 456 | - |
| Serous carcinoma | yes | 0 | 0 | 0 | 0 | 0 | 0 | 0 | 0 | 0 | 0 | 0 | **0** | **0.0** |
| | no | 9 | 11 | 105 | 12 | 25 | 8 | 21 | 145 | 35 | 7 | 34 | 412 | - |
| Clear cell carcinoma | yes | 0 | 0 | 0 | 0 | 0 | 0 | 0 | 0 | 0 | 0 | 0 | **0** | **0.0** |
| | no | 4 | 0 | 62 | 18 | 14 | 2 | 6 | 36 | 16 | 0 | 5 | 163 | - |
| Carcinosarcoma | yes | 0 | 0 | 0 | 0 | 0 | 0 | 0 | 0 | 0 | 0 | 0 | **0** | **0.0** |
| | no | 0 | 0 | 0 | 0 | 24 | 3 | 8 | 13 | 12 | 0 | 38 | 98 | - |
| Un-dedifferentiated | yes | 0 | 0 | 0 | 0 | 0 | 0 | 0 | 0 | 0 | 0 | 0 | **0** | **0.0** |
| | no | 0 | 0 | 14 | 0 | 9 | 3 | 3 | 17 | 6 | 0 | 0 | 52 | - |
| Other | yes | 0 | 0 | 0 | 0 | 0 | 0 | 0 | 0 | 0 | 0 | 0 | **0** | **0.0** |
| | no | 5 | 1 | 37 | 0 | 29 | 24 | 1 | 9 | 8 | 107 | 6 | 227 | - |
| **LVSI** | | | | | | | | | | | | | | |
| Present | yes | 3 | 5 | 111 | 15 | 8 | 9 | 7 | 10 | 28 | 0 | 23 | **219** | **21.2** |
| | no | 23 | 15 | 278 | 40 | 45 | 0 | 15 | 45 | 20 | 0 | 22 | 503 | - |
| Focal or absent | yes | 97 | 85 | 66 | 9 | 29 | 141 | 5 | 54 | 92 | 0 | 41 | **619** | **59.9** |
| | no | 440 | 288 | 205 | 31 | 173 | 19 | 18 | 300 | 63 | 0 | 57 | 1594 | - |
| Unknown | yes | 14 | 4 | 0 | 0 | 0 | 4 | 2 | 0 | 9 | 164 | 1 | **198** | **18.9** |
| | no | 137 | 30 | 0 | 21 | 16 | 23 | 8 | 3 | 10 | 114 | 6 | 368 | - |
| **2009 FIGO stage** | | | | | | | | | | | | | | |
| IA | yes | 44 | 13 | 8 | 3 | 0 | 22 | 0 | 11 | 68 | 111* | 20 | **300** | **66.1*** |
| | no | 250 | 58 | 70 | 8 | 28 | 22 | 0 | 163 | 27 | 66* | 30 | 722 | - |
| IB | yes | 70 | 80 | 27 | 6 | 7 | 129 | 0 | 15 | 33 | - | 16 | **383** | - |
| | no | 350 | 271 | 90 | 25 | 67 | 20 | 0 | 81 | 16 | - | 9 | 929 | - |
| II | yes | 0 | 0 | 48 | 5 | 11 | 0 | 0 | 2 | 6 | 18 | 10 | **100** | **9.7** |
| | no | 0 | 2 | 122 | 16 | 71 | 0 | 0 | 27 | 8 | 14 | 14 | 274 | - |
| IIIA | yes | 0 | 0 | 29 | 4 | 11 | 0 | 0 | 1 | 7 | 29* | 5 | **86** | **20.5*** |
| | no | 0 | 2 | 54 | 14 | 41 | 0 | 0 | 5 | 7 | 27* | 8 | 158 | - |
| IIIB | yes | 0 | 1 | 10 | 0 | 5 | 0 | 0 | 4 | 1 | - | 1 | **22** | - |
| | no | 0 | 0 | 32 | 4 | 14 | 0 | 0 | 10 | 2 | - | 0 | 62 | - |
| IIIC | yes | 0 | 0 | 55 | 5 | 3 | 0 | 0 | 21 | 10 | - | 10 | **104** | - |
| | no | 0 | 0 | 115 | 14 | 12 | 0 | 0 | 40 | 7 | - | 20 | 208 | - |
| IV | yes | 0 | 0 | 0 | 1 | 0 | 0 | 14 | 10 | 1 | 6 | 0 | **32** | **3.1** |
| | no | 0 | 0 | 0 | 10 | 1 | 0 | 41 | 22 | 3 | 7 | 1 | 85 | - |
| Unkown | yes | 0 | 0 | 0 | 0 | 0 | 3 | 0 | 0 | 3 | - | 3 | **9** | **0.6** |
| | no | 0 | 0 | 0 | 1 | 0 | 0 | 0 | 0 | 23 | - | 3 | 27 | - |
| **Molecular class** | | | | | | | | | | | | | | |
| *POLE* mut | yes | 9 | 6 | 19 | 3 | 2 | 13 | 0 | 5 | 4 | 0 | 9 | **70** | **6.8** |
| | no | 33 | 18 | 32 | 13 | 14 | 2 | 0 | 34 | 1 | 0 | 1 | 148 | - |
| MMRd | yes | 22 | 19 | 57 | 6 | 8 | 49 | 7 | 29 | 43 | 0 | 28 | **268** | **25.9** |
| | no | 115 | 91 | 82 | 14 | 63 | 5 | 3 | 80 | 5 | 0 | 8 | 466 | - |
| p53abn | yes | 8 | 3 | 16 | 3 | 7 | 3 | 1 | 17 | 7 | 0 | 6 | **71** | **6.9** |
| | no | 32 | 27 | 83 | 31 | 62 | 9 | 33 | 184 | 48 | 0 | 67 | 576 | - |
| NSMP | yes | 51 | 61 | 65 | 12 | 20 | 79 | 5 | 13 | 66 | 0 | 19 | **391** | **37.9** |
| | no | 214 | 171 | 57 | 31 | 70 | 1 | 4 | 50 | 17 | 0 | 8 | 623 | - |
| Unkown | yes | 24 | 5 | 20 | 0 | 0 | 10 | 1 | 0 | 9 | 164 | 3 | **236** | **22.6** |
| | no | 206 | 26 | 229 | 3 | 25 | 25 | 1 | 0 | 22 | 114 | 1 | 652 | - |

**Table S7 Detailed composition by cohort of the supervised training dataset for the task Low-grade (grade 1-2) Endometrioid versus High-grade (grade 3) Endometrioid.** The inclusion variable represents the number of patients (with one WSI for each) included in the supervised training task. The percentage column is computed over the total number of included patients. For the UMCG cohort, only 2009 FIGO stage I, II, and III was known (as opposed to the subtype IA, IB, and IIIA, IIIB, IIIC), hence stage proportion numbers denoted with * represent pooled stage I or pooled stage III. *POLE*mut = *POLE* mutant; MMRd = Mismatch repair deficient; p53abn = p53 abnormal; NSMP = No specific molecular profile; LVSI = Lymphovascular space invasion.

#### *POLE*mut versus wildtype task

| | inclusion | PORTEC-1 | PORTEC-2 | PORTEC-3 | transPORTEC | MST-I | MST-II | DOMEC | DANISH | LUMC | UMCG | SOUTH-AFRICA | Total | % of included |
|---|---|---|---|---|---|---|---|---|---|---|---|---|---|---|
| **Number of patients** | | | | | | | | | | | | | | |
| | yes | 92 | 95 | 236 | 28 | 50 | 147 | 44 | 20 | 184 | 0 | 130 | **1026** | - |
| | no | 622 | 332 | 424 | 88 | 221 | 49 | 11 | 392 | 38 | 278 | 20 | 2475 | - |
| **Age: median** | | | | | | | | | | | | | | |
| | yes | 67 | 69 | 62 | 66 | 64 | 68 | - | 71 | 68 | - | 65 | **67** | - |
| | no | 66 | 69 | 62 | 68 | 70 | 65 | - | 69 | 64 | 64 | 65 | **66** | - |
| **Histotype** | | | | | | | | | | | | | | |
| Endometrioid grade 1-2 | yes | 72 | 80 | 92 | 4 | 21 | 111 | 8 | 0 | 94 | 0 | 26 | **508** | **49.5** |
| | no | 529 | 294 | 165 | 14 | 80 | 19 | 2 | 0 | 19 | 156 | 5 | 1283 | - |
| Endometrioid grade 3 | yes | 18 | 9 | 65 | 20 | 16 | 19 | 5 | 8 | 26 | 0 | 26 | **212** | **20.7** |
| | no | 77 | 32 | 120 | 48 | 53 | 3 | 1 | 184 | 6 | 8 | 3 | 535 | - |
| Serous carcinoma | yes | 0 | 6 | 38 | 3 | 2 | 7 | 16 | 11 | 27 | 0 | 30 | **140** | **13.6** |
| | no | 9 | 5 | 67 | 9 | 23 | 1 | 5 | 134 | 8 | 7 | 4 | 272 | - |
| Clear cell carcinoma | yes | 0 | 0 | 24 | 1 | 2 | 2 | 6 | 0 | 14 | 0 | 3 | **52** | **5.1** |
| | no | 4 | 0 | 38 | 17 | 12 | 0 | 0 | 36 | 2 | 0 | 2 | 111 | - |
| Carcinosarcoma | yes | 0 | 0 | 0 | 0 | 3 | 3 | 6 | 0 | 11 | 0 | 32 | **55** | **5.4** |
| | no | 0 | 0 | 0 | 0 | 21 | 0 | 2 | 13 | 1 | 0 | 6 | 43 | - |
| Un-dedifferentiated | yes | 0 | 0 | 5 | 0 | 2 | 3 | 3 | 1 | 5 | 0 | 0 | **19** | **1.9** |
| | no | 0 | 0 | 9 | 0 | 7 | 0 | 0 | 16 | 1 | 0 | 0 | 33 | - |
| Other | yes | 2 | 0 | 12 | 0 | 4 | 2 | 0 | 0 | 7 | 0 | 13 | **40** | **3.9** |
| | no | 3 | 1 | 25 | 0 | 25 | 26 | 1 | 9 | 1 | 107 | 0 | 198 | - |
| **LVSI** | | | | | | | | | | | | | | |
| Present | yes | 3 | 5 | 149 | 18 | 11 | 7 | 21 | 1 | 44 | 0 | 40 | **299** | **29.1** |
| | no | 23 | 15 | 240 | 37 | 42 | 2 | 1 | 54 | 4 | 0 | 5 | 423 | - |
| Focal or absent | yes | 82 | 86 | 87 | 10 | 39 | 138 | 20 | 19 | 126 | 0 | 84 | **691** | **67.3** |
| | no | 455 | 287 | 184 | 30 | 163 | 22 | 3 | 335 | 29 | 0 | 14 | 1522 | - |
| Unknown | yes | 7 | 4 | 0 | 0 | 0 | 2 | 3 | 0 | 14 | 0 | 6 | **36** | **3.5** |
| | no | 144 | 30 | 0 | 21 | 16 | 25 | 7 | 3 | 5 | 278 | 1 | 530 | - |
| **2009 FIGO stage** | | | | | | | | | | | | | | |
| IA | yes | 38 | 15 | 31 | 3 | 2 | 27 | 0 | 7 | 79 | 0* | 44 | **246** | **24.3** |
| | no | 256 | 56 | 47 | 8 | 26 | 17 | 0 | 167 | 16 | 177* | 6 | 776 | - |
| IB | yes | 54 | 79 | 42 | 6 | 10 | 117 | 0 | 8 | 39 | - | 22 | **377** | **36.7** |
| | no | 366 | 272 | 75 | 25 | 64 | 32 | 0 | 88 | 10 | - | 3 | 935 | - |
| II | yes | 0 | 0 | 55 | 5 | 11 | 0 | 0 | 0 | 13 | 0 | 22 | **106** | **10.3** |
| | no | 0 | 2 | 115 | 16 | 71 | 0 | 0 | 29 | 1 | 32 | 2 | 268 | - |
| IIIA | yes | 0 | 0 | 29 | 5 | 16 | 0 | 0 | 0 | 14 | 0* | 10 | **74** | **7.2** |
| | no | 0 | 2 | 54 | 13 | 36 | 0 | 0 | 6 | 0 | 56* | 3 | 170 | - |
| IIIB | yes | 0 | 1 | 14 | 0 | 8 | 0 | 0 | 0 | 3 | - | 1 | **27** | **2.6** |
| | no | 0 | 0 | 28 | 4 | 11 | 0 | 0 | 14 | 0 | - | 0 | 57 | - |
| IIIC | yes | 0 | 0 | 65 | 8 | 3 | 0 | 0 | 4 | 15 | - | 28 | **123** | **12.0** |
| | no | 0 | 0 | 105 | 11 | 12 | 0 | 0 | 57 | 2 | - | 2 | 189 | - |
| IV | yes | 0 | 0 | 0 | 1 | 0 | 0 | 44 | 1 | 2 | 0 | 0 | **48** | **4.7** |
| | no | 0 | 0 | 0 | 10 | 1 | 0 | 11 | 31 | 2 | 13 | 1 | 69 | - |
| Unkown | yes | 0 | 0 | 0 | 0 | 0 | 3 | 0 | 0 | 19 | 0 | 3 | **25** | **2.4** |
| | no | 0 | 0 | 0 | 1 | 0 | 0 | 0 | 0 | 7 | 0 | 3 | 11 | - |
| **Molecular class** | | | | | | | | | | | | | | |
| *POLE* mut | yes | 10 | 7 | 29 | 3 | 4 | 14 | 0 | 1 | 5 | 0 | 8 | **81** | **7.9** |
| | no | 33 | 17 | 22 | 13 | 12 | 0 | 0 | 37 | 0 | 0 | 0 | 134 | - |
| MMRd | yes | 22 | 20 | 77 | 6 | 10 | 48 | 7 | 9 | 48 | 0 | 35 | **282** | **27.5** |
| | no | 115 | 90 | 62 | 14 | 61 | 5 | 3 | 101 | 0 | 0 | 1 | 452 | - |
| p53abn | yes | 9 | 4 | 56 | 6 | 15 | 11 | 28 | 8 | 54 | 0 | 64 | **255** | **24.9** |
| | no | 31 | 26 | 43 | 28 | 54 | 0 | 6 | 182 | 1 | 0 | 9 | 380 | - |
| NSMP | yes | 51 | 64 | 74 | 13 | 21 | 74 | 9 | 2 | 77 | 0 | 23 | **408** | **39.8** |
| | no | 213 | 168 | 48 | 30 | 69 | 10 | 0 | 72 | 6 | 0 | 6 | 622 | - |
| Unkown | yes | 0 | 0 | 0 | 0 | 0 | 0 | 0 | 0 | 0 | 0 | 0 | **0** | **0.0** |
| | no | 230 | 31 | 249 | 3 | 25 | 34 | 2 | 0 | 31 | 278 | 4 | 887 | - |

**Table S8 Detailed composition by cohort of the supervised training dataset for the task *POLE*mut versus wildtype.** The inclusion variable represents the number of patients (with one WSI for each) included in the supervised training task. The percentage column is computed over the total number of included patients. For the UMCG cohort, only 2009 FIGO stage I, II, and III was known (as opposed to the subtype IA, IB, and IIIA, IIIB, IIIC), hence stage proportion numbers denoted with * represent pooled stage I or pooled stage III. *POLE*mut = *POLE* mutant; MMRd = Mismatch repair deficient; p53abn = p53 abnormal; NSMP = No specific molecular profile; LVSI = Lymphovascular space invasion.

#### MMRd versus MMRp task

| | inclusion | PORTEC-1 | PORTEC-2 | PORTEC-3 | transPORTEC | MST-I | MST-II | DOMEC | DANISH | LUMC | UMCG | SOUTH-AFRICA | Total | % of included |
|---|---|---|---|---|---|---|---|---|---|---|---|---|---|---|
| **Number of patients** | | | | | | | | | | | | | | |
| | yes | 120 | 100 | 268 | 28 | 52 | 171 | 45 | 20 | 191 | 0 | 142 | **1137** | - |
| | no | 594 | 327 | 392 | 88 | 219 | 25 | 10 | 392 | 31 | 278 | 8 | 2364 | - |
| **Age: median** | | | | | | | | | | | | | | |
| | yes | 67 | 69 | 62 | 66 | 64 | 68 | - | 71 | 68 | - | 65 | **67** | - |
| | no | 66 | 70 | 62 | 68 | 70 | 65 | - | 69 | 62 | 64 | 68 | **67** | - |
| **Histotype** | | | | | | | | | | | | | | |
| Endometrioid grade 1-2 | yes | 95 | 85 | 104 | 4 | 21 | 129 | 9 | 0 | 97 | 0 | 30 | **574** | **50.6** |
| | no | 506 | 289 | 153 | 14 | 80 | 1 | 1 | 0 | 16 | 156 | 1 | 1217 | - |
| Endometrioid grade 3 | yes | 19 | 9 | 73 | 20 | 16 | 21 | 5 | 8 | 27 | 0 | 29 | **227** | **20.0** |
| | no | 76 | 32 | 112 | 48 | 53 | 1 | 1 | 184 | 5 | 8 | 0 | 520 | - |
| Serous carcinoma | yes | 1 | 6 | 44 | 3 | 2 | 8 | 16 | 11 | 29 | 0 | 33 | **153** | **13.5** |
| | no | 8 | 5 | 61 | 9 | 23 | 0 | 5 | 134 | 6 | 7 | 1 | 259 | - |
| Clear cell carcinoma | yes | 2 | 0 | 27 | 1 | 2 | 2 | 6 | 0 | 15 | 0 | 4 | **59** | **5.2** |
| | no | 2 | 0 | 35 | 17 | 12 | 0 | 0 | 36 | 1 | 0 | 1 | 104 | - |
| Carcinosarcoma | yes | 0 | 0 | 0 | 0 | 3 | 3 | 6 | 0 | 11 | 0 | 33 | **56** | **4.9** |
| | no | 0 | 0 | 0 | 0 | 21 | 0 | 2 | 13 | 1 | 0 | 5 | 42 | - |
| Un-dedifferentiated | yes | 0 | 0 | 8 | 0 | 2 | 3 | 3 | 1 | 5 | 0 | 0 | **22** | **1.9** |
| | no | 0 | 0 | 6 | 0 | 7 | 0 | 0 | 16 | 1 | 0 | 0 | 30 | - |
| Other | yes | 3 | 0 | 12 | 0 | 6 | 5 | 0 | 0 | 7 | 0 | 13 | **46** | **3.9** |
| | no | 2 | 1 | 25 | 0 | 23 | 23 | 1 | 9 | 1 | 107 | 0 | 192 | - |
| **LVSI** | | | | | | | | | | | | | | |
| Present | yes | 4 | 6 | 168 | 18 | 12 | 9 | 21 | 1 | 44 | 0 | 45 | **328** | **28.9** |
| | no | 22 | 14 | 221 | 37 | 41 | 0 | 1 | 54 | 4 | 0 | 0 | 394 | - |
| Focal or absent | yes | 102 | 90 | 100 | 10 | 39 | 158 | 21 | 19 | 132 | 0 | 90 | **761** | **67.0** |
| | no | 435 | 283 | 171 | 30 | 163 | 2 | 2 | 335 | 23 | 0 | 8 | 1452 | - |
| Unknown | yes | 14 | 4 | 0 | 0 | 1 | 4 | 3 | 0 | 15 | 0 | 7 | **48** | **4.1** |
| | no | 137 | 30 | 0 | 21 | 15 | 23 | 7 | 3 | 4 | 278 | 0 | 518 | - |
| **2009 FIGO stage** | | | | | | | | | | | | | | |
| IA | yes | 47 | 16 | 35 | 3 | 2 | 36 | 0 | 7 | 82 | 0* | 47 | **275** | **24.2** |
| | no | 247 | 55 | 43 | 8 | 26 | 8 | 0 | 167 | 13 | 177* | 3 | 747 | - |
| IB | yes | 73 | 83 | 47 | 6 | 11 | 133 | 0 | 8 | 41 | - | 24 | **426** | 66.0 |
| | no | 347 | 268 | 70 | 25 | 63 | 16 | 0 | 88 | 8 | - | 1 | 886 | - |
| II | yes | 0 | 0 | 64 | 5 | 12 | 0 | 0 | 0 | 14 | 0 | 23 | **118** | **10.4** |
| | no | 0 | 2 | 106 | 16 | 70 | 0 | 0 | 29 | 0 | 32 | 1 | 256 | - |
| IIIA | yes | 0 | 0 | 32 | 5 | 16 | 0 | 0 | 0 | 14 | 0* | 10 | **77** | **6.8** |
| | no | 0 | 2 | 51 | 13 | 36 | 0 | 0 | 6 | 0 | 56* | 3 | 167 | - |
| IIIB | yes | 0 | 1 | 15 | 0 | 8 | 0 | 0 | 0 | 3 | - | 1 | **28** | **2.5** |
| | no | 0 | 0 | 27 | 4 | 11 | 0 | 0 | 14 | 0 | - | 0 | 56 | - |
| IIIC | yes | 0 | 0 | 75 | 8 | 3 | 0 | 0 | 4 | 16 | - | 30 | **136** | **12.0** |
| | no | 0 | 0 | 95 | 11 | 12 | 0 | 0 | 57 | 1 | - | 0 | 176 | - |
| IV | yes | 0 | 0 | 0 | 1 | 0 | 0 | 45 | 1 | 2 | 0 | 1 | **50** | **4.4** |
| | no | 0 | 0 | 0 | 10 | 1 | 0 | 10 | 31 | 2 | 13 | 0 | 67 | - |
| Unkown | yes | 0 | 0 | 0 | 0 | 0 | 2 | 0 | 0 | 19 | 0 | 6 | **27** | **2.4** |
| | no | 0 | 0 | 0 | 1 | 0 | 1 | 0 | 0 | 7 | 0 | 0 | 9 | - |
| **Molecular class** | | | | | | | | | | | | | | |
| *POLE* mut | yes | 10 | 7 | 29 | 3 | 4 | 14 | 0 | 1 | 5 | 0 | 8 | **81** | **7.9** |
| | no | 33 | 17 | 22 | 13 | 12 | 0 | 0 | 37 | 0 | 0 | 0 | 134 | - |
| MMRd | yes | 22 | 20 | 77 | 6 | 10 | 54 | 7 | 9 | 48 | 0 | 36 | **289** | **25.5** |
| | no | 115 | 90 | 62 | 14 | 61 | 0 | 3 | 101 | 0 | 0 | 0 | 446 | - |
| p53abn | yes | 9 | 4 | 56 | 6 | 15 | 12 | 28 | 8 | 54 | 0 | 68 | **260** | **22.9** |
| | no | 31 | 26 | 43 | 28 | 54 | 0 | 6 | 182 | 1 | 0 | 5 | 376 | - |
| NSMP | yes | 52 | 64 | 74 | 13 | 21 | 80 | 9 | 2 | 77 | 0 | 26 | **418** | **36.7** |
| | no | 213 | 168 | 48 | 30 | 69 | 2 | 0 | 72 | 6 | 0 | 3 | 611 | - |
| Unkown | yes | 27 | 5 | 32 | 0 | 2 | 11 | 1 | 0 | 7 | 0 | 4 | **89** | **7.7** |
| | no | 202 | 26 | 217 | 3 | 23 | 23 | 1 | 0 | 24 | 278 | 0 | 797 | - |

**Table S9 Detailed composition by cohort of the supervised training dataset for the task MMRd versus MMRp.** The inclusion variable represents the number of patients (with one WSI for each) included in the supervised training task. The percentage column is computed over the total number of included patients. For the UMCG cohort, only 2009 FIGO stage I, II, and III was known (as opposed to the subtype IA, IB, and IIIA, IIIB, IIIC), hence stage proportion numbers denoted with * represent pooled stage I or pooled stage III. For this task, all patients with a positive IHC-MMR (MMRd) are counted as positive cases. *POLE*mut = *POLE* mutant; MMRd = Mismatch repair deficient; p53abn = p53 abnormal; NSMP = No specific molecular profile; LVSI = Lymphovascular space invasion.

#### p53abn versus wildtype task

| | inclusion | PORTEC-1 | PORTEC-2 | PORTEC-3 | transPORTEC | MST-I | MST-II | DOMEC | DANISH | LUMC | UMCG | SOUTH-AFRICA | Total | % of included |
|---|---|---|---|---|---|---|---|---|---|---|---|---|---|---|
| **Number of patients** | | | | | | | | | | | | | | |
| | yes | 120 | 100 | 268 | 28 | 52 | 171 | 45 | 20 | 191 | 0 | 142 | 1137 | - |
| | no | 594 | 327 | 392 | 88 | 219 | 25 | 10 | 392 | 31 | 278 | 8 | 2364 | - |
| **Age: median** | | | | | | | | | | | | | | |
| | yes | 67 | 69 | 62 | 66 | 64 | 68 | - | 71 | 68 | - | 65 | 67 | - |
| | no | 66 | 70 | 62 | 68 | 70 | 65 | - | 69 | 62 | 64 | 69 | 67 | - |
| **Histotype** | | | | | | | | | | | | | | |
| Endometrioid grade 1-2 | yes | 95 | 85 | 104 | 4 | 21 | 129 | 9 | 0 | 97 | 0 | 30 | 574 | 50.6 |
| | no | 506 | 289 | 153 | 14 | 80 | 1 | 1 | 0 | 16 | 156 | 1 | 1217 | - |
| Endometrioid grade 3 | yes | 19 | 9 | 73 | 20 | 16 | 21 | 5 | 8 | 27 | 0 | 29 | 227 | 20.0 |
| | no | 76 | 32 | 112 | 48 | 53 | 1 | 1 | 184 | 5 | 8 | 0 | 520 | - |
| Serous carcinoma | yes | 1 | 6 | 44 | 3 | 2 | 8 | 16 | 11 | 29 | 0 | 33 | 153 | 13.5 |
| | no | 8 | 5 | 61 | 9 | 23 | 0 | 5 | 134 | 6 | 7 | 1 | 259 | - |
| Clear cell carcinoma | yes | 2 | 0 | 27 | 1 | 2 | 2 | 6 | 0 | 15 | 0 | 4 | 59 | 5.2 |
| | no | 2 | 0 | 35 | 17 | 12 | 0 | 0 | 36 | 1 | 0 | 1 | 104 | - |
| Carcinosarcoma | yes | 0 | 0 | 0 | 0 | 3 | 3 | 6 | 0 | 11 | 0 | 33 | 56 | 4.9 |
| | no | 0 | 0 | 0 | 0 | 21 | 0 | 2 | 13 | 1 | 0 | 5 | 42 | - |
| Un-dedifferentiated | yes | 0 | 0 | 8 | 0 | 2 | 3 | 3 | 1 | 5 | 0 | 0 | 22 | 1.9 |
| | no | 0 | 0 | 6 | 0 | 7 | 0 | 0 | 16 | 1 | 0 | 0 | 30 | - |
| Other | yes | 3 | 0 | 12 | 0 | 6 | 5 | 0 | 0 | 7 | 0 | 13 | 46 | 3.9 |
| | no | 2 | 1 | 25 | 0 | 23 | 23 | 1 | 9 | 1 | 107 | 0 | 192 | - |
| **LVSI** | | | | | | | | | | | | | | |
| Present | yes | 4 | 6 | 168 | 18 | 12 | 9 | 21 | 1 | 44 | 0 | 45 | 328 | 28.9 |
| | no | 22 | 14 | 221 | 37 | 41 | 0 | 1 | 54 | 4 | 0 | 0 | 394 | - |
| Focal or absent | yes | 102 | 90 | 100 | 10 | 39 | 158 | 21 | 19 | 132 | 0 | 90 | 761 | 67.0 |
| | no | 435 | 283 | 171 | 30 | 163 | 25 | 2 | 335 | 23 | 0 | 8 | 1475 | - |
| Unknown | yes | 14 | 4 | 0 | 0 | 1 | 4 | 3 | 0 | 15 | 0 | 7 | 48 | 4.1 |
| | no | 137 | 30 | 0 | 21 | 15 | 0 | 7 | 3 | 4 | 278 | 0 | 495 | - |
| **2009 FIGO stage** | | | | | | | | | | | | | | |
| IA | yes | 47 | 16 | 35 | 3 | 2 | 36 | 0 | 7 | 82 | 0* | 47 | 275 | 24.2 |
| | no | 247 | 55 | 43 | 8 | 26 | 8 | 0 | 167 | 13 | 177* | 3 | 747 | - |
| IB | yes | 73 | 83 | 47 | 6 | 11 | 133 | 0 | 8 | 41 | - | 24 | 426 | 37.5 |
| | no | 347 | 268 | 70 | 25 | 63 | 16 | 0 | 88 | 8 | - | 1 | 886 | - |
| II | yes | 0 | 0 | 64 | 5 | 12 | 0 | 0 | 0 | 14 | 0 | 23 | 118 | 10.4 |
| | no | 0 | 2 | 106 | 16 | 70 | 0 | 0 | 29 | 0 | 32 | 1 | 256 | - |
| IIIA | yes | 0 | 0 | 32 | 5 | 16 | 0 | 0 | 0 | 14 | 0* | 10 | 77 | 6.8 |
| | no | 0 | 2 | 51 | 13 | 36 | 0 | 0 | 6 | 0 | 56* | 3 | 167 | - |
| IIIB | yes | 0 | 1 | 15 | 0 | 8 | 0 | 0 | 0 | 3 | - | 1 | 28 | 2.5 |
| | no | 0 | 0 | 27 | 4 | 11 | 0 | 0 | 14 | 0 | - | 0 | 56 | - |
| IIIC | yes | 0 | 0 | 75 | 8 | 3 | 0 | 0 | 4 | 16 | - | 30 | 136 | 12.0 |
| | no | 0 | 0 | 95 | 11 | 12 | 0 | 0 | 57 | 1 | - | 0 | 176 | - |
| IV | yes | 0 | 0 | 0 | 1 | 0 | 0 | 45 | 1 | 2 | 0 | 1 | 50 | 4.4 |
| | no | 0 | 0 | 0 | 10 | 1 | 0 | 10 | 31 | 2 | 13 | 0 | 67 | - |
| Unkown | yes | 0 | 0 | 0 | 0 | 0 | 2 | 0 | 0 | 19 | 0 | 6 | 27 | 2.4 |
| | no | 0 | 0 | 0 | 1 | 0 | 1 | 0 | 0 | 7 | 0 | 0 | 9 | - |
| **Molecular class** | | | | | | | | | | | | | | |
| *POLE*mut | yes | 10 | 7 | 29 | 3 | 4 | 14 | 0 | 1 | 5 | 0 | 8 | 81 | 7.9 |
| | no | 33 | 17 | 22 | 13 | 12 | 0 | 0 | 37 | 0 | 0 | 0 | 134 | - |
| MMRd | yes | 22 | 20 | 77 | 6 | 10 | 54 | 7 | 9 | 48 | 0 | 36 | 289 | 25.5 |
| | no | 115 | 90 | 62 | 14 | 61 | 0 | 3 | 101 | 0 | 0 | 0 | 446 | - |
| p53abn | yes | 9 | 4 | 56 | 6 | 15 | 12 | 28 | 8 | 54 | 0 | 38 | 230 | 20.3 |
| | no | 31 | 26 | 43 | 28 | 54 | 0 | 6 | 182 | 1 | 0 | 5 | 376 | - |
| NSMP | yes | 52 | 64 | 74 | 13 | 21 | 80 | 9 | 2 | 77 | 0 | 24 | 416 | 36.7 |
| | no | 213 | 168 | 48 | 30 | 69 | 0 | 0 | 72 | 6 | 0 | 3 | 609 | - |
| Unkown | yes | 27 | 5 | 32 | 0 | 2 | 11 | 1 | 0 | 7 | 0 | 36 | 121 | 10.3 |
| | no | 202 | 26 | 217 | 3 | 23 | 25 | 1 | 0 | 24 | 278 | 0 | 799 | - |

**Table S10 Detailed composition by cohort of the supervised training dataset for the task p53abn versus wildtype.** The inclusion variable represents the number of patients (with one WSI for each) included in the supervised training task. The percentage column is computed over the total number of included patients. For the UMCG cohort, only 2009 FIGO stage I, II, and III was known (as opposed to the subtype IA, IB, and IIIA, IIIB, IIIC), hence stage proportion numbers denoted with * represent pooled stage I or pooled stage III. For this task, all patients with a positive IHC-p53 are counted as positive cases. *POLE*mut = *POLE* mutant; MMRd = Mismatch repair deficient; p53abn = p53 abnormal; NSMP = No specific molecular profile; LVSI = Lymphovascular space invasion.

#### ER positive versus negative task

| | inclusion | PORTEC-1 | PORTEC-2 | PORTEC-3 | ansPORTE | MST-I | MST-II | DOMEC | DANISH | LUMC | UMCG | SOUTH-AFRICA | Total | % of included |
|---|---|---|---|---|---|---|---|---|---|---|---|---|---|---|
| **Number of patients** | | | | | | | | | | | | | | |
| | yes | 84 | 98 | 223 | 3 | 52 | 172 | 45 | 142 | 191 | 0 | 142 | **1152** | - |
| | no | 630 | 329 | 437 | 113 | 219 | 24 | 10 | 270 | 31 | 278 | 8 | 2349 | - |
| **Age: median** | | | | | | | | | | | | | | |
| | yes | 68 | 69 | 63 | 78 | 64 | 68 | - | 69 | 68 | - | 65 | **68** | - |
| | no | 66 | 69 | 62 | 68 | 70 | 65 | - | 70 | 62 | 64 | 69 | **67** | - |
| **Histotype** | | | | | | | | | | | | | | |
| Endometrioid grade 1-2 | yes | 67 | 84 | 88 | 0 | 21 | 130 | 9 | 0 | 97 | 0 | 30 | **526** | **45.7** |
| | no | 534 | 290 | 169 | 18 | 80 | 0 | 1 | 0 | 16 | 156 | 1 | 1265 | - |
| Endometrioid grade 3 | yes | 15 | 9 | 60 | 3 | 16 | 21 | 5 | 64 | 27 | 0 | 29 | **249** | **21.7** |
| | no | 80 | 32 | 125 | 65 | 53 | 1 | 1 | 128 | 5 | 8 | 0 | 498 | - |
| Serous carcinoma | yes | 0 | 5 | 39 | 0 | 2 | 8 | 16 | 54 | 29 | 0 | 33 | **186** | **16.2** |
| | no | 9 | 6 | 66 | 12 | 23 | 0 | 5 | 91 | 6 | 7 | 1 | 226 | - |
| Clear cell carcinoma | yes | 0 | 0 | 21 | 0 | 2 | 2 | 6 | 13 | 15 | 0 | 4 | **63** | **5.5** |
| | no | 4 | 0 | 41 | 18 | 12 | 0 | 0 | 23 | 1 | 0 | 1 | 100 | - |
| Carcinosarcoma | yes | 0 | 0 | 0 | 0 | 3 | 3 | 6 | 4 | 11 | 0 | 33 | **60** | **5.2** |
| | no | 0 | 0 | 0 | 0 | 21 | 0 | 2 | 9 | 1 | 0 | 5 | 38 | - |
| Un-dedifferentiated | yes | 0 | 0 | 4 | 0 | 2 | 3 | 3 | 5 | 5 | 0 | 0 | **22** | **1.9** |
| | no | 0 | 0 | 10 | 0 | 7 | 0 | 0 | 12 | 1 | 0 | 0 | 30 | - |
| Other | yes | 2 | 0 | 11 | 0 | 6 | 5 | 0 | 2 | 7 | 0 | 13 | **46** | **3.8** |
| | no | 3 | 1 | 26 | 0 | 23 | 23 | 1 | 7 | 1 | 107 | 0 | 192 | - |
| **LVSI** | | | | | | | | | | | | | | |
| Present | yes | 3 | 5 | 142 | 0 | 12 | 9 | 21 | 24 | 44 | 0 | 45 | **305** | **26.5** |
| | no | 23 | 15 | 247 | 55 | 41 | 0 | 1 | 31 | 4 | 0 | 0 | 417 | - |
| Focal or absent | yes | 76 | 89 | 81 | 3 | 39 | 159 | 21 | 118 | 132 | 0 | 90 | **808** | **70.3** |
| | no | 461 | 284 | 190 | 37 | 163 | 1 | 2 | 236 | 23 | 0 | 8 | 1405 | - |
| Unknown | yes | 5 | 4 | 0 | 0 | 1 | 4 | 3 | 0 | 15 | 0 | 7 | **39** | **3.2** |
| | no | 146 | 30 | 0 | 21 | 15 | 23 | 7 | 3 | 4 | 278 | 0 | 527 | - |
| **2009 FIGO stage** | | | | | | | | | | | | | | |
| IA | yes | 35 | 15 | 26 | 0 | 2 | 34 | 0 | 31 | 82 | 0* | 47 | **272** | **23.7** |
| | no | 259 | 56 | 52 | 11 | 26 | 10 | 0 | 143 | 13 | 177* | 3 | 750 | - |
| IB | yes | 49 | 82 | 39 | 2 | 11 | 136 | 0 | 27 | 41 | - | 24 | **411** | **35.7** |
| | no | 371 | 269 | 78 | 29 | 63 | 13 | 0 | 69 | 8 | - | 1 | 901 | - |
| II | yes | 0 | 0 | 54 | 1 | 12 | 0 | 0 | 7 | 14 | 0 | 23 | **111** | **9.7** |
| | no | 0 | 2 | 116 | 20 | 70 | 0 | 0 | 22 | 0 | 32 | 1 | 263 | - |
| IIIA | yes | 0 | 0 | 27 | 0 | 16 | 0 | 0 | 4 | 14 | 0* | 10 | **71** | **6.2** |
| | no | 0 | 2 | 56 | 18 | 36 | 0 | 0 | 2 | 0 | 56* | 3 | 173 | - |
| IIIB | yes | 0 | 1 | 15 | 0 | 8 | 0 | 0 | 5 | 3 | - | 1 | **33** | **2.9** |
| | no | 0 | 0 | 27 | 4 | 11 | 0 | 0 | 9 | 0 | - | 0 | 51 | - |
| IIIC | yes | 0 | 0 | 62 | 0 | 3 | 0 | 0 | 42 | 16 | - | 30 | **153** | **13.3** |
| | no | 0 | 0 | 108 | 19 | 12 | 0 | 0 | 19 | 1 | - | 0 | 159 | - |
| IV | yes | 0 | 0 | 0 | 0 | 0 | 0 | 45 | 26 | 2 | 0 | 1 | **74** | **6.4** |
| | no | 0 | 0 | 0 | 11 | 1 | 0 | 10 | 6 | 2 | 13 | 0 | 43 | - |
| Unkown | yes | 0 | 0 | 0 | 0 | 0 | 2 | 0 | 0 | 19 | 0 | 6 | **27** | **2.3** |
| | no | 0 | 0 | 0 | 1 | 0 | 1 | 0 | 0 | 7 | 0 | 0 | 9 | - |
| **Molecular class** | | | | | | | | | | | | | | |
| *POLE* mut | yes | 9 | 7 | 25 | 1 | 4 | 14 | 0 | 8 | 5 | 0 | 8 | **81** | **7.1** |
| | no | 34 | 17 | 26 | 15 | 12 | 0 | 0 | 30 | 0 | 0 | 0 | 134 | - |
| MMRd | yes | 18 | 19 | 69 | 0 | 10 | 54 | 7 | 37 | 48 | 0 | 36 | **298** | **25.9** |
| | no | 119 | 91 | 70 | 20 | 61 | 0 | 3 | 73 | 0 | 0 | 0 | 437 | - |
| p53abn | yes | 9 | 4 | 54 | 1 | 15 | 12 | 28 | 70 | 54 | 0 | 68 | **315** | **27.3** |
| | no | 31 | 26 | 45 | 33 | 54 | 0 | 6 | 120 | 1 | 0 | 5 | 321 | - |
| NSMP | yes | 48 | 64 | 70 | 1 | 21 | 80 | 9 | 27 | 77 | 0 | 24 | **421** | **36.5** |
| | no | 216 | 168 | 52 | 42 | 69 | 0 | 0 | 47 | 6 | 0 | 3 | 603 | - |
| Unkown | yes | 0 | 4 | 5 | 0 | 2 | 12 | 1 | 0 | 7 | 0 | 6 | **37** | **3.2** |
| | no | 230 | 27 | 244 | 3 | 23 | 24 | 1 | 0 | 24 | 278 | 0 | 854 | - |

**Table S11 Detailed composition by cohort of the supervised training dataset for the task ER positive versus negative.** The inclusion variable represents the number of patients (with one WSI for each) included in the supervised training task. The percentage column is computed over the total number of included patients. For the UMCG cohort, only 2009 FIGO stage I, II, and III was known (as opposed to the subtype IA, IB, and IIIA, IIIB, IIIC), hence stage proportion numbers denoted with * represent pooled stage I or pooled stage III. *POLE*mut = *POLE* mutant; MMRd = Mismatch repair deficient; p53abn = p53 abnormal; NSMP = No specific molecular profile; LVSI = Lymphovascular space invasion.

#### Distant recurrence task

| | inclusion | PORTEC-1 | PORTEC-2 | PORTEC-3 | transPORTEC | MST-I | MST-II | DOMEC | DANISH | LUMC | UMCG | SOUTH-AFRICA | Total | % of included |
|---|---|---|---|---|---|---|---|---|---|---|---|---|---|---|
| **Number of patients** | | | | | | | | | | | | | | |
| | yes | 120 | 100 | 43 | 13 | 35 | 171 | 0 | 47 | 152 | 167 | 0 | 848 | - |
| | no | 594 | 327 | 617 | 103 | 236 | 25 | 55 | 365 | 70 | 111 | 150 | 2653 | - |
| **Follow-up: median** | | | | | | | | | | | | | | |
| | yes | 10.7 | 10.0 | 5.2 | 3.0 | 3.3 | 9.3 | - | 6.4 | 2.8 | 5.3 | - | 5.3 | - |
| | no | 10.4 | 9.49 | 5.0 | 2.0 | 5.6 | 11.1 | - | 6.2 | 2.3 | 4.2 | - | 5.6 | - |
| **Age: median** | | | | | | | | | | | | | | |
| | yes | 67 | 69 | 63 | 71 | 65 | 68 | - | 70 | 68 | 66 | 65 | 68 | |
| | no | 66 | 70 | 62 | 67 | 70 | 64 | - | 69 | 66 | 61 | - | 66 | - |
| **Histotype** | | | | | | | | | | | | | | |
| Endometrioid grade 1-2 | yes | 95 | 85 | 19 | 1 | 14 | 130 | 0 | 0 | 90 | 148 | 0 | 582 | 68.6 |
| | no | 506 | 289 | 238 | 17 | 87 | 0 | 10 | 0 | 23 | 8 | 31 | 1209 | - |
| Endometrioid grade 3 | yes | 19 | 9 | 8 | 10 | 11 | 21 | 0 | 25 | 22 | 7 | 0 | 132 | 15.6 |
| | no | 76 | 32 | 177 | 58 | 58 | 1 | 6 | 167 | 10 | 1 | 29 | 615 | - |
| Serous carcinoma | yes | 1 | 6 | 8 | 2 | 2 | 8 | 0 | 14 | 17 | 7 | 0 | 65 | 7.6 |
| | no | 8 | 5 | 97 | 10 | 23 | 0 | 21 | 131 | 18 | 0 | 34 | 347 | - |
| Clear cell carcinoma | yes | 2 | 0 | 7 | 0 | 2 | 2 | 0 | 6 | 12 | 0 | 0 | 31 | 3.7 |
| | no | 2 | 0 | 55 | 18 | 12 | 0 | 6 | 30 | 4 | 0 | 5 | 132 | - |
| Carcinosarcoma | yes | 0 | 0 | 0 | 0 | 2 | 3 | 0 | 1 | 3 | 0 | 0 | 9 | 1.1 |
| | no | 0 | 0 | 0 | 0 | 22 | 0 | 8 | 12 | 9 | 0 | 38 | 89 | - |
| Un-dedifferentiated | yes | 0 | 0 | 1 | 0 | 1 | 3 | 0 | 1 | 3 | 0 | 0 | 9 | 1.1 |
| | no | 0 | 0 | 13 | 0 | 8 | 0 | 3 | 16 | 3 | 0 | 0 | 43 | - |
| Other | yes | 3 | 0 | 0 | 0 | 3 | 4 | 0 | 0 | 5 | 5 | 0 | 20 | 2.4 |
| | no | 2 | 1 | 37 | 0 | 26 | 24 | 1 | 9 | 3 | 102 | 13 | 218 | - |
| **LVSI** | | | | | | | | | | | | | | |
| Present | yes | 4 | 6 | 23 | 6 | 7 | 9 | 0 | 2 | 28 | 0 | 0 | 85 | 10.0 |
| | no | 22 | 14 | 366 | 49 | 46 | 0 | 22 | 53 | 20 | 0 | 45 | 637 | - |
| Focal or absent | yes | 102 | 90 | 20 | 7 | 27 | 159 | 0 | 45 | 117 | 0 | 0 | 567 | 66.9 |
| | no | 435 | 283 | 251 | 33 | 175 | 1 | 23 | 309 | 38 | 0 | 98 | 1646 | - |
| Unknown | yes | 14 | 4 | 0 | 0 | 1 | 3 | 0 | 0 | 7 | 167 | 0 | 196 | 23.1 |
| | no | 137 | 30 | 0 | 21 | 15 | 24 | 10 | 3 | 12 | 111 | 7 | 370 | - |
| **2009 FIGO stage** | | | | | | | | | | | | | | |
| IA | yes | 47 | 16 | 5 | 1 | 2 | 36 | 0 | 24 | 77 | 117* | 0 | 325 | 81.3‡ |
| | no | 247 | 55 | 73 | 10 | 26 | 10 | 0 | 150 | 18 | 60* | 50 | 639 | - |
| IB | yes | 73 | 83 | 6 | 4 | 9 | 135 | 0 | 12 | 42 | - | 0 | 364 | - |
| | no | 347 | 268 | 111 | 27 | 65 | 15 | 0 | 84 | 7 | - | 25 | 949 | - |
| II | yes | 0 | 0 | 10 | 4 | 7 | 0 | 0 | 3 | 14 | 19 | 0 | 57 | 6.7 |
| | no | 0 | 2 | 160 | 17 | 75 | 0 | 0 | 26 | 0 | 13 | 24 | 317 | - |
| IIIA | yes | 0 | 0 | 6 | 2 | 11 | 0 | 0 | 2 | 8 | 31* | 0 | 60 | 12.0‡ |
| | no | 0 | 2 | 77 | 16 | 41 | 0 | 0 | 4 | 6 | 25* | 13 | 184 | - |
| IIIB | yes | 0 | 1 | 1 | 0 | 4 | 0 | 0 | 3 | 1 | - | 0 | 10 | - |
| | no | 0 | 0 | 41 | 4 | 15 | 0 | 0 | 11 | 2 | - | 1 | 74 | - |
| IIIC | yes | 0 | 0 | 15 | 2 | 2 | 0 | 0 | 3 | 10 | - | 0 | 32 | - |
| | no | 0 | 0 | 155 | 17 | 13 | 0 | 0 | 58 | 7 | - | 30 | 280 | - |
| IV | yes | 0 | 0 | 0 | 0 | 0 | 0 | 0 | 0 | 0 | 0 | 0 | 0 | 0.0 |
| | no | 0 | 0 | 0 | 11 | 1 | 0 | 55 | 32 | 4 | 13 | 1 | 117 | - |
| Unkown | yes | 0 | 0 | 0 | 0 | 0 | 0 | 0 | 0 | 0 | 0 | 0 | 0 | 0.0 |
| | no | 0 | 0 | 0 | 1 | 0 | 0 | 0 | 0 | 26 | 26 | 26 | 79 | - |
| **Molecular class** | | | | | | | | | | | | | | |
| *POLE*mut | yes | 9 | 7 | 6 | 2 | 2 | 15 | 0 | 2 | 5 | 0 | 0 | 48 | 5.6 |
| | no | 34 | 17 | 45 | 14 | 14 | 0 | 0 | 36 | 0 | 0 | 10 | 170 | - |
| MMRd | yes | 22 | 20 | 11 | 1 | 7 | 54 | 0 | 14 | 40 | 0 | 0 | 169 | 19.9 |
| | no | 115 | 90 | 128 | 19 | 64 | 0 | 10 | 96 | 8 | 0 | 36 | 566 | - |
| p53abn | yes | 9 | 4 | 8 | 3 | 9 | 12 | 0 | 20 | 36 | 0 | 0 | 101 | 11.9 |
| | no | 31 | 26 | 91 | 31 | 60 | 0 | 34 | 170 | 19 | 0 | 73 | 535 | - |
| NSMP | yes | 52 | 64 | 11 | 7 | 16 | 80 | 0 | 11 | 66 | 0 | 0 | 307 | 36.2 |
| | no | 212 | 168 | 111 | 36 | 74 | 0 | 9 | 63 | 17 | 0 | 27 | 717 | - |
| Unkown | yes | 28 | 5 | 7 | 0 | 1 | 10 | 0 | 0 | 5 | 167 | 0 | 223 | 26.3 |
| | no | 202 | 26 | 242 | 3 | 24 | 25 | 2 | 0 | 26 | 111 | 4 | 665 | - |
| **Adjuvant treatment (%)** | | | | | | | | | | | | | | |
| None | yes | 67 | 0 | 0 | 1 | 0 | 2 | 0 | 44 | 76 | 0 | 0 | 190 | 22.4 |
| | no | 302 | 3 | 0 | 9 | 0 | 0 | 0 | 227 | 11 | 0 | 0 | 552 | - |
| Radiotherapy | yes | 53 | 100 | 43 | 12 | 35 | 169 | 0 | 3 | 76 | 167 | 0 | 658 | 77.6 |
| | no | 292 | 324 | 290 | 54 | 209 | 25 | 0 | 32 | 4 | 0 | 0 | 1230 | - |
| Chemotherapy**/Chemoradiatio | yes | 0 | 0 | 0 | 0 | 0 | 0 | 0 | 0 | 0 | 0 | 0 | 0 | 0.0 |
| | no | 0 | 0 | 327 | 16 | 22 | 0 | 55 | 106 | 37 | 3 | 0 | 566 | - |
| Unknown | yes | 0 | 0 | 0 | 0 | 0 | 0 | 0 | 0 | 0 | 0 | 0 | 0 | 0.0 |
| | no | 0 | 0 | 0 | 24 | 5 | 0 | 0 | 0 | 18 | 108 | 150 | 305 | - |

**Table S12 Detailed composition by cohort of the supervised training dataset for the prognostic task of distant recurrence risk prediction.** The inclusion variable represents the number of patients (with one WSI for each) included in the supervised training task. The percentage column is computed over the total number of included patients. For the UMCG cohort, only 2009 FIGO stage I, II, and III was known (as opposed to the subtype IA, IB, and IIIA, IIIB, IIIC), hence stage proportion numbers denoted with * represent pooled stage I or pooled stage III. Patients with a 2009 FIGO stage IV and those who received chemotherapy, chemoradiation (**or hormone therapy) were excluded. *POLE*mut = *POLE* mutant; MMRd =

Mismatch repair deficient; p53abn = p53 abnormal; NSMP = No specific molecular profile; LVSI = Lymphovascular space invasion.
